# Temporal multi-omic analysis of COVID-19 in end-stage kidney disease

**DOI:** 10.1101/2024.06.20.24309228

**Authors:** Emily Stephenson, Erin Macdonald-Dunlop, Lisa M Dratva, Rik G.H. Lindeboom, Zewen Kelvin Tuong, Win Min Tun, Norzawani B Buang, Stephane Ballereau, Mia Cabantaus, Ana Peñalver, Elena Prigmore, John R Ferdinand, Benjamin J Stewart, Jack Gisby, Talat Malik, Candice L Clarke, Nicholas Medjeral-Thomas, Maria Prendecki, Stephen McAdoo, Anais Portet, Michelle Willicombe, Eleanor Sandhu, Matthew C. Pickering, Marina Botto, Sarah A. Teichmann, Muzlifah Haniffa, Menna R. Clatworthy, David C. Thomas, James E. Peters

## Abstract

**Summary:** Patients with end-stage kidney disease (ESKD) are at high risk of severe COVID-19. We performed longitudinal single cell multi-omic immune profiling of ESKD patients with COVID- 19, sampled during two waves of the pandemic. Uniquely, for a subset of patients, we obtained samples before and during acute infection, allowing intra-individual comparison. Using single-cell transcriptome, surface proteome and immunoreceptor sequencing of 580,040 high-quality cells, derived from 187 longitudinal samples from 61 patients, we demonstrate widespread changes following infection. We identified gene expression signatures of severity, with the majority of pathways differentiating mild from severe disease in B cells and monocytes. For example, gene expression of *PLAC8*, a receptor known to modulate SARS-CoV-2 entry to cells, was a marker of severity in CD14+ monocytes. Longitudinal profiling demonstrated distinct temporal molecular trajectories in severe versus mild disease, including type 1 and type 2 interferon signalling, *MHC* gene expression and, in B cells, a proliferative signature (*KRAS* and *MYC*). Evaluation of clonal T cell dynamics showed that the fastest expanding clones were significantly enriched in known SARS-CoV-2 specific sequences and shared across multiple patients. Our analyses revealed novel TCR clones likely reactive to SARS-CoV-2. Finally, we identified a population of transcriptionally distinct monocytes that emerged in peripheral blood following glucocorticoid treatment. Overall, our data delineate the temporal dynamics of the immune response in COVID-19 in a high-risk population and provide a valuable open-access resource.

## Introduction

COVID-19, caused by the SARS-CoV-2 virus, displays marked clinical heterogeneity, varying from minimal symptoms to fatal disease. This variation in outcome is not random; severe or fatal COVID-19 disproportionately affects certain strata of the population. Demographic risk factors for severe COVID-19 include older age, male sex, and non-white ethnicity. Underlying medical conditions also impact the risk of severe COVID-19. End-stage kidney disease (ESKD) is one of the strongest risk factors for severe COVID-19, with a UK population-scale study estimating a hazard ratio for death of 3.69^1^. While vaccines have been highly effective in reducing morbidity and mortality from SARS-CoV-2, novel viral variants of concern (VoC) continue to emerge and ESKD patients remain at elevated risk of hospitalisation and death. There is, therefore, a need for research and therapeutic efforts focusing on ESKD patients and other high-risk groups.

A central feature of the pathophysiology of severe COVID-19 is an excessive host inflammatory response leading to tissue injury. Autopsies revealed an accumulation of activated immune cells but little or no active virus^2^. Severe disease is characterised by excess circulating monocytes, neutrophils and myeloid progenitors and elevated pro-inflammatory cytokines and chemokines, which contribute to endothelial damage and the formation of microthrombi. The importance of the host immune response is further underscored by the efficacy of therapies targeting inflammation. Glucocorticoids, which have pleiotropic effects on inflammatory pathways, and targeted inhibition of the IL6 signalling pathway, both reduce mortality in COVID-19^3,4,5^. Drugs targeting the JAK-STAT pathway also appear promising^6^.

ESKD is defined as irreversible loss of renal function with a glomerular filtration rate <15 mls/min/1.73m^2^ and is fatal without dialysis or transplantation. In addition to loss of glomerular filtration, ESKD is a systemic disease associated with profound changes in hormonal, cardiovascular and haematopoietic function^7^. Such disturbances of normal physiology are also associated with changes in immune function, and ESKD patients have both increased susceptibility to infection and impaired vaccination responses^8^. Conversely, despite evidence of impaired specific adaptive immune responses, ESKD is also characterised by a chronic pro- inflammatory state^7^.

Thus, patients with ESKD may be at high risk of complications of SARS-CoV-2 due to their preponderance of cardiometabolic risk factors, as well as both impaired immunity and pro-inflammatory state secondary to uraemia. An outstanding question is whether patients with ESKD mount a qualitatively different immune response to SARS-CoV-2 that drives their susceptibility to severe COVID-19. Furthermore, the need to attend medical facilities for regular haemodialysis, regardless of infection with SARS-CoV-2, provides a unique opportunity to evaluate the temporal dynamics of the host immune response through serial sample collection in both the inpatient and outpatient setting.

Here, we longitudinally profile the evolving immune cellular landscape driven by COVID-19 infection in ESKD patients, allowing us to produce single-cell resolution multi-omic temporal trajectories in the context of renal failure. Uniquely, we collected samples from the same set of individuals before and during SARS-CoV-2 infection. Our data provide a rich resource for examining the complex longitudinal dynamics of COVID-19 infection in the context of an important and clinically vulnerable patient group.

## Results

### Longitudinal immune cell profiling in ESKD patients with COVID-19

To investigate the dynamics of the immune cellular landscape in ESKD patients with COVID- 19, we performed longitudinal blood sampling and profiling of peripheral blood mononuclear cells (PBMC) in ESKD patients. Patients were recruited from a single centre in London, UK, during two distinct waves of COVID-19. The first cohort (‘2020 Cohort’) (n=22) were recruited in April-May 2020, during the initial phase of the pandemic and before the advent of vaccination. This cohort consisted of COVID-19 positive ESKD patients, including both inpatients and outpatients, with a spectrum of illness severity from mild to critical (**Fig. 1A, Supp.Table 1**). Following COVID-19 diagnosis, serial blood sampling was performed over the course of the illness (**Fig 1B**). In addition, we recruited COVID-19 negative ESKD patients to provide an appropriate control group. This group was well-matched in terms of age, sex, and ethnicity (**Supp.Table 1**).

**Fig. 1:**
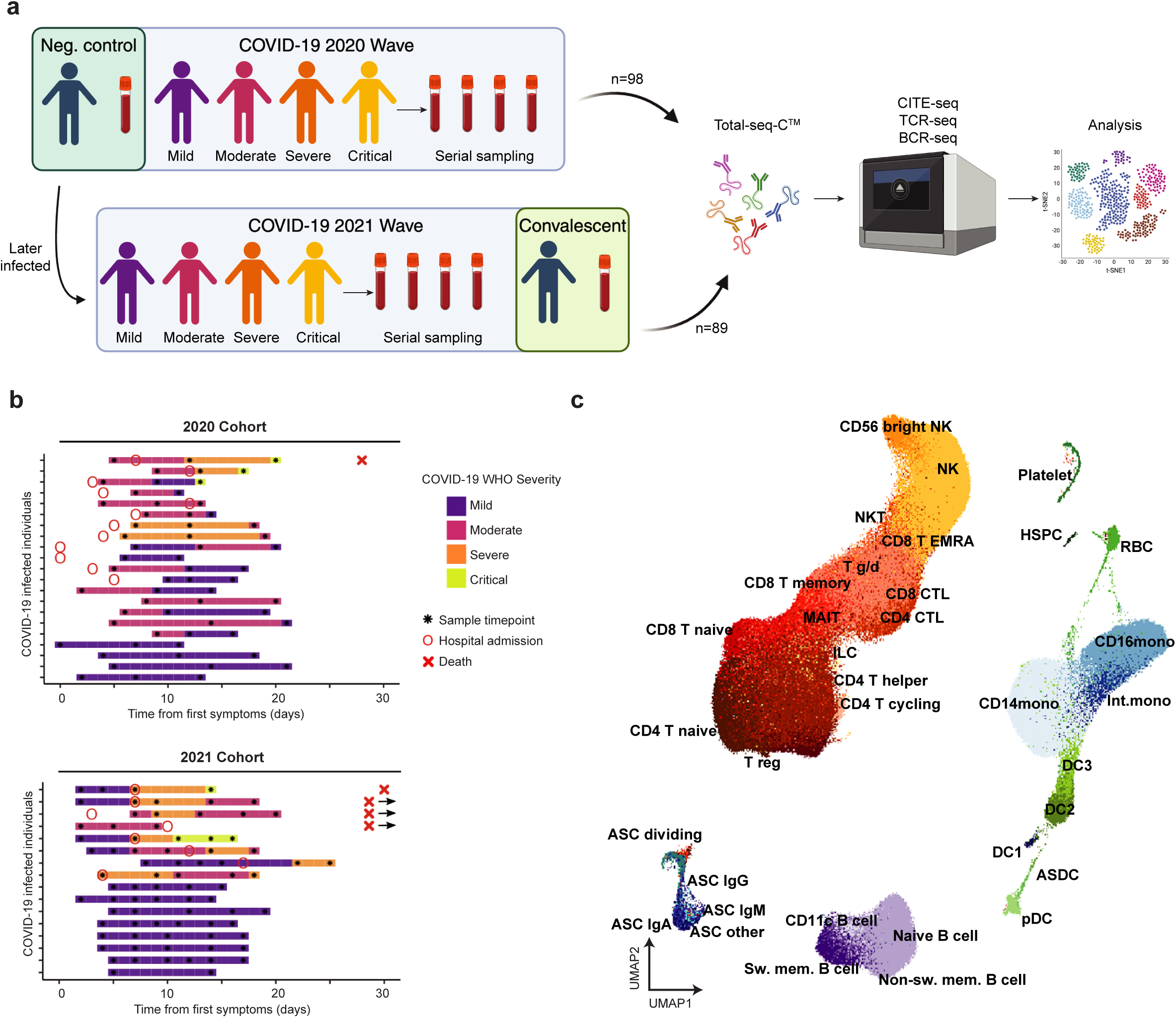
Study Overview. **A)** Schematic of the study design showing the recruitment of both cohorts and how their samples were processed in the laboratory then analysed. Neg. control = COVID-19 negative ESKD patient. Figure created using Biorender.com. **B)** Timing of blood sampling in relation to illness onset. Colours indicate the COVID-19 severity over time. ‘X’ with an adjacent arrow indicates death during the hospital admission occurring at >30 days. **C)** UMAP showing the detailed annotations of B cells, myeloid and progenitors and T cells, respectively. ASDC = Axl Siglec dendritic cell, MAIT = mucosal-associated invariant T cell, ASC = antibody secreting cell, sw. mem. = switched memory, CTL = cytotoxic T lymphocyte, T g/d = gamma delta T cell, EMRA = terminally differentiated effector memory T cell, ILC = innate lymphoid cell, mono = monocyte, int. = intermediate.

The second cohort (‘2021 Cohort’) (n=16) consisted of ESKD patients with COVID-19, sampled between January-March 2021 (when the alpha variant was the predominant variant in the UK). These patients were recruited as part of the COVID-19 negative control group in the 2020 Cohort. Again, serial blood sampling was performed during the acute illness. Thus, for the 2021 Cohort we had matched samples from pre-infection and during acute COVID-19, enabling intra-individual analysis. In addition, for a subset of the 2021 Cohort we collected a convalescent sample approximately 2 months after infection (n=10). (**Fig. 1A-B**).

To provide a comprehensive yet granular assessment of cellular and molecular changes, we used a single-cell multi-omic approach, performing Cellular Indexing of Transcriptomes and Epitopes by sequencing (CITE-seq) of PBMC samples with matched T cell receptor sequencing (TCR-seq) and B cell receptor sequencing (BCR-seq) (**Fig. 1A, Supp.Table 2**). To exclude low quality cells, we removed those with less than 200 genes, more than 10% mitochondrial reads and fewer than 1000 UMIs. We performed genotype-based demultiplexing of pooled samples (Methods) and removed cells with genotypes not fully resolvable. The full dataset consisted of 580,040 cells, representing 187 longitudinal samples from 61 patients (median of 3 samples per patient).

For initial cell type annotation, we separated the data analyses into three broad categories of cell types: i) B cells, ii) T cells and innate lymphocytes, and iii) myeloid and non-immune hematopoietic cells (**Fig. 1C**). Data were integrated per patient to account for technical artefacts (Methods). Using semi-automatic cell type annotations with CellTypist^9^, previously published COVID-19 reference atlases^10,11^, and canonical marker genes, we were able to identify 39 cell types (**Fig. 1C, Supp.** Fig. 1A-D). These comprised known subtypes of monocytes (classical CD14+ mono, non-classical CD16+ mono and intermediate CD14+ CD16+ mono) and dendritic cells (DCs), plus sub-populations displaying an interferon- stimulated signature^11^ and complement expressing CD16+ monocytes that we have described previously (**Fig. 1C, Supp.** Fig. 1B)^10^. Within the B cell compartment, we leveraged the availability of paired single-cell BCR-seq and CITE-seq data to detect 9 sub-populations. (**Fig. 1C, Supp.** Fig. 1C). Similarly, using TCR-seq and CITE-seq data, we were able to recover 15 clusters encompassing T cells, natural killer (NK) cells and innate-like lymphocytes (ILC) (**Fig. 1C, Supp.** Fig. 1D).

### Altered cellular and transcriptomic profiles in ESKD patients with COVID-19

To understand the changes in peripheral immune cellular proportions in COVID-19, we compared samples from COVID-19 positive ESKD patients to those from COVID-19 negative ESKD patients. We first compared samples during the first week of COVID-19 (week 1) to COVID-19 negative samples. Analysis using broad cell type annotations showed a significant decrease in the relative abundance of the total monocyte population in COVID-19 positive versus COVID-19 negative ESKD patients (**Fig. 2A, Supp.Table 3).** More granular cell-type categorisation revealed that samples taken during week 1 from COVID-19 positive patients had a significantly lower relative abundance of all monocyte subsets including classical (CD14 mono), non-classical (CD16 mono) and intermediate CD14+CD16+ (Int. mono) monocytes (**Fig. 2B-D, Supp.Table 3)**. Relative abundances of CD8+ memory T cells and CD4+ CTL and DC3 were reduced while those of naive B cells increased (**Fig. 2E-H, Supp.Table 3)**. There were no significant changes in cell abundance between samples taken in the second week of COVID-19 compared to the first week.

**Fig. 2:**
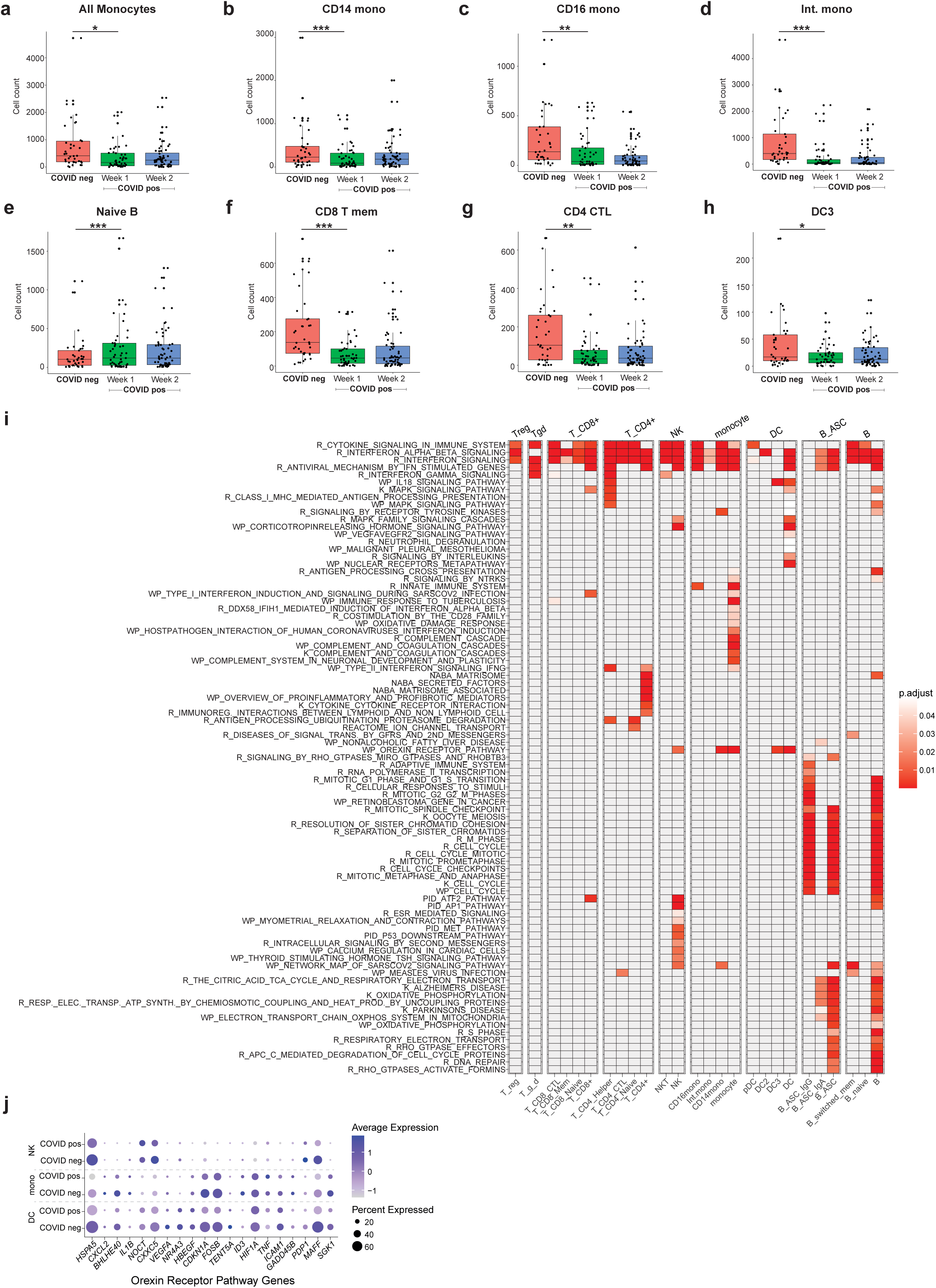
**Cell type abundance and DEG/pathway analysis for positive cases versus negative controls**. **A-H)** Bar charts displaying relative numbers of cells that significantly changed in abundance in week 1 of COVID-19 infection compared to a control group of COVID-19 negative ESKD patients. *P ≤ 0.05, **P ≤ 0.01, ***P ≤ 0.001. **I)** Heatmap of gene expression pathways significantly (FDR &lt;0.05) associated with COVID-19 positivity. Cell labels above plot denote broad cell annotations and labels below denote more granular cell annotations. R = reactome, WP = wiki pathways, K = KEGG, NABA = Alexandra Naba and PID = pathway interaction database. **J)** Dot plot displaying the expression of the leading-edge subset of genes that contributed to the term “Orexin Receptor Pathway’’ for COVID positive and negative patients. Mono = monocytes.

Next, to assess immune cell transcriptomic changes in COVID-19 in ESKD patients, we performed differential gene expression analysis within each cell type, comparing samples from COVID-19 positive and COVID-19 negative patients (**Supp.Table 4**). To identify the biological pathways implicated by these differentially expressed genes, we performed gene set enrichment analysis (**Supp.Table 5**). We observed widespread transcriptomic changes between COVID-19 positive and negative samples across numerous cell types. In the 2020 Cohort, the most consistent finding was an enrichment of interferon alpha and beta response pathways across a broad range of innate and adaptive immune cells (**Fig. 2I, Supp.Table 5**).

B cells exhibited the greatest number of significantly enriched pathway terms, totalling 240 pathways (**Supp.Table 5**). Many of these contain genes that are involved in the cell cycle and DNA repair, likely reflecting the strong B cell proliferative response involved in initiating adaptive immunity to SARS-CoV-2. Similarly, expression of genes relating to protein translation and post-translational modification were up-regulated, likely relating to the generation of an antibody response. Many of the enriched pathways were also noted in B cell antibody-secreting cells (B-ASC). Examining other cell types, we identified 19 enriched pathways in monocytes, 17 in NK cells, 15 in dendritic cells, 8 in CD4+ T cells, 7 in CD8+ T cells (**Fig. 2I, Supp.Table 5**).

Some pathways were significantly enriched between COVID-19 positive and COVID-19- negative ESKD samples across multiple cell types. For example, we observed a significant negative enrichment of the “Orexin Receptor Pathway’’ across multiple innate immune cell types, including monocytes (adjusted P 2.36x10^-5^), dendritic cells (adjusted P 2.01x10^-8^), and NK cells (adjusted P 0.014) (**Supp.Table 5**). The leading-edge subset of genes that contributed to this term included *SGK1, GADD45B, MAFF, PDP1, ICAM1, TENT5A, HIF1A, TNF, ID3, CDKN1A, FOSB, HBEGF, NR4A3, VEGFA, BHLHE40, CXXC5, NOCT, IL1B,*

*CXCL2* and *HSPA5* (**Fig. 2J**), many of which are cellular stress response genes. Analysis of

monocyte subsets using more fine-grained annotation, revealed enrichment of the pathway specifically in classical CD14 monocytes, and not in intermediate and non-classical CD16 monocytes, suggesting the former was the source of the signal in monocytes. In the smaller 2021 Cohort, where we had paired pre-infection and infection samples from the same individuals, we replicated the findings of significant enrichment of the “Orexin Receptor Pathway’’ in CD14+ CD16- monocytes and NK cells, but not in dendritic cells (**Supp.Table 5**).

Overall, our findings comparing COVID-19 positive and negative samples from ESKD patients are broadly similar to those described previously in other more general patient populations.

### Immune cell transcriptomic correlates of COVID-19 severity in ESKD patients

We next assessed molecular and cellular changes associated with COVID-19 severity at the time of blood sampling, categorised as mild, moderate, severe, or critical (Methods). Analysis of cell type numbers comparing samples taken from patients at the time of severe or critical disease (hereafter ‘severe/critical’, n=56) to those taken at the time of mild or moderate disease (hereafter ‘mild/moderate’, n=84) revealed that the relative abundance of dividing B- ASC cells was increased in the severe/critical group (**Fig. 3A, Supp.Table 6**).

**Fig. 3:**
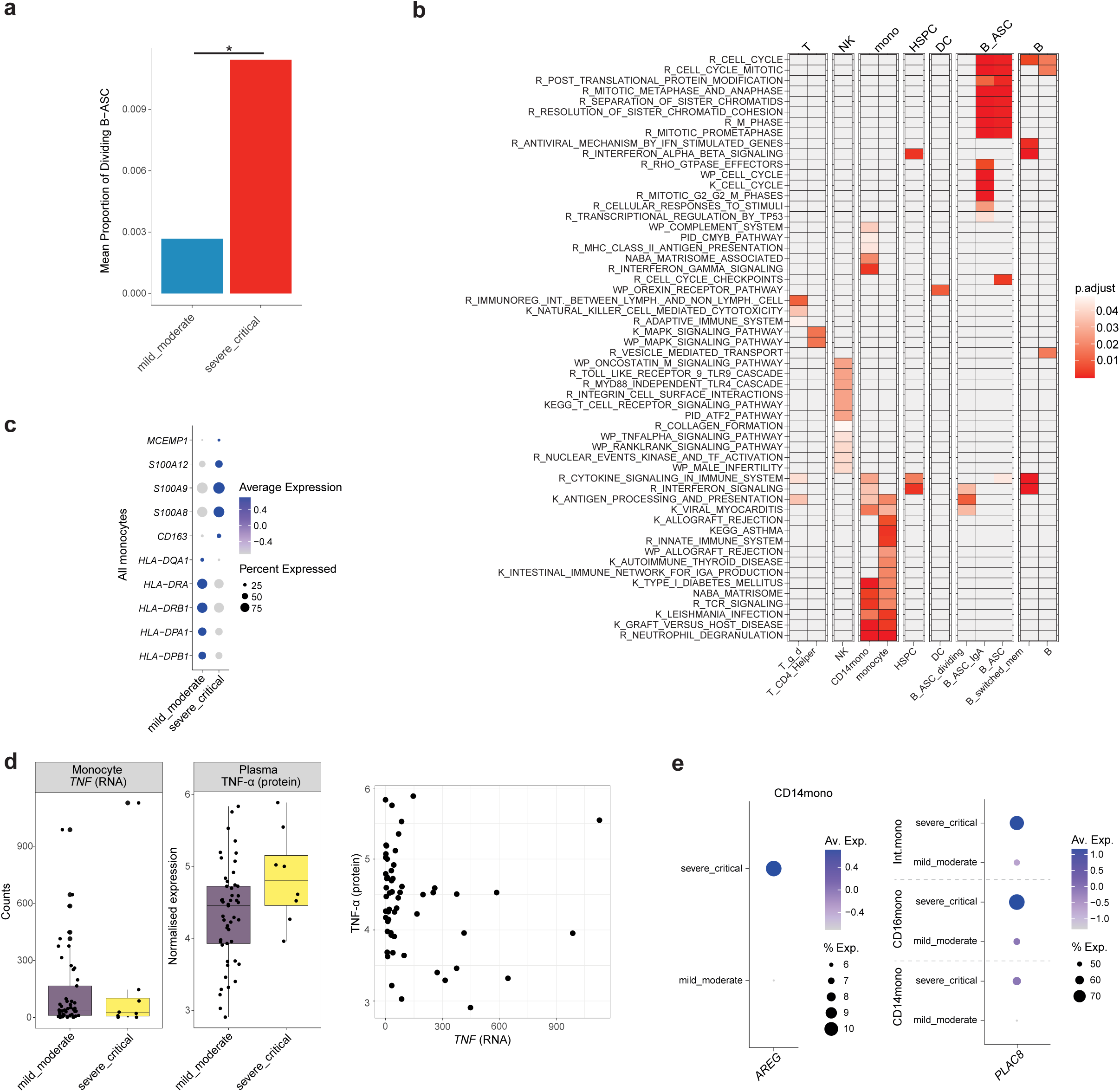
**Cell type abundance and DEG/pathway analysis for severity of positive cases**. **A)** Bar chart displaying the relative abundance of dividing antibody secreting B cells for mild and moderate patients compared to severe and critical patients. P = 0.017 **B)** Heatmap of pathways significantly (FDR &lt;0.05) associated with COVID-19 severity. Cell labels above plot denote broad cell annotations and labels below denote more granular cell annotations. R = reactome, WP = wiki pathways, K = KEGG and PID = pathway interaction database. **C)** Dot plot displaying the expression of differentially expressed genes relating to severity in all monocyte populations. **D)** Left: Boxplots of *TNF* RNA counts in monocytes from the scRNA-seq dataset and normalised plasma TNF-ɑ protein abundance measured with Olink immunoassays (n=57 samples from 21 individuals with both RNA and plasma protein levels measured). Right: correlation between *TNF* gene expression and TNF-ɑ plasma protein levels. (Pearson r -0.15) **E)** Left: dot plot displaying gene expression of *AREG* in CD14+ monocytes stratified according to COVID-19 severity at the time of sampling. Right: dot plot displaying gene expression of *PLAC8* in CD14+, CD16+ and intermediate monocytes split, again stratified according to COVID-19 severity.

We then performed differential gene expression within each cell type, again comparing samples taken at the time of severe/critical COVID-19 to mild/moderate COVID-19 (**Supp.Table 7**). Taking genes significantly associated with disease severity, we identified the corresponding biological pathways through enrichment analysis (**Fig. 3B, Supp.Table 8**). Where significant enrichment of pathways was identified within a cell type, we then used a more granular cell type annotation to delineate the source of the signal. We identified 86 pathways that differentiated mild/moderate from severe/critical disease. The majority of these were in the B cell (35) or monocyte (29) compartment. 11 pathways were associated with severity in NK cells and 5 in gamma delta cells with only 2 enriched pathways in CD4+ helper T cells with none in CD8+ T cells. Similarly, at the single gene level, 205 genes distinguished mild/moderate from severe/critical disease. 125 of these were in all monocytes or CD14+CD16- classical monocytes and 21 were in B cell subsets **(Supp.Table 8)**.

In B cells, “Antibody secreting cells” and “Antibody secreting cells that produce IgA”, numerous pathways relating to cell division were enriched in severe/critical disease. This is likely to represent increased activation of the adaptive immune response in severe/critical disease and the initiation of a response to drive neutralising antibody production. In dividing antibody secreted cells (B_ASC_dividing) and switched memory B cells, increased interferon signalling pathways also distinguished severe/critical from mild/moderate disease (**Supp.Table 8**).

In monocytes, differential gene expression analysis between mild/moderate and severe/critical samples revealed significant enrichment of numerous pathways, including the KEGG “Asthma” and “Graft Versus Host Disease”, “Leishmania Infection” and “Allograft Rejection” pathway terms (**Supp.Table 8**). Many genes in these pathways were downregulated in severe/critical relative to mild/moderate disease and their high enrichment scores are driven, in part, by the high representation of *HLA* genes in the pathways. This is likely to reflect the downregulation of MHC molecules on antigen presenting cells in severe COVID-19 that has been previously reported. We observed down-regulation of *HLA-DPB1*, *HLA-DPA1*, *HLA- DRB1*, *HLA-DRA* and *HLA-DQA1* in all monocytes (**Fig. 3C**). This was accompanied by down- regulation of *CD163* as noted in other studies. We also noted up-regulation of genes previously associated with severity such as *S100A8*, *S100A9*, *S100A12* and *MCEMP1* **(Fig. 3C)**^12^. In all monocyte subsets, the most differentially expressed gene between mild/moderate and severe/critical samples was *TNF*, encoding TNF-α (p-value 6.4 x10^-116^) **(Supp.Table 7)**. Unexpectedly, given its pro-inflammatory effects, *TNF* gene expression was lower in severe/critical disease. We hypothesised that this might be as a result of negative feedback from elevated TNF-ɑ at the protein level. We therefore compared monocyte *TNF* gene expression levels to protein levels of plasma TNF-ɑ measured using Olink immunoassays in the same set of samples. This revealed higher plasma TNF-ɑ protein in samples taken at the time of severe/critical disease. Correlation analysis between plasma TNF-ɑ protein monocyte *TNF* gene expression revealed a weak negative correlation (Pearson r -0.15), demonstrating an uncoupling of plasma protein and gene expression levels **(Fig. 3D)**.

Given the importance of monocytes in the host immune contribution to COVID-19 severity^13^, we performed a deeper analysis of specific subsets. In classical CD14+ monocytes, there was enrichment for the Reactome “MHC Class II Antigen Presentation”, KEGG “Type I Diabetes Mellitus” and “Graft Versus Host Disease” and Reactome “Interferon Gamma Signalling” in severe disease **(Supp.Table 8)**. Many of these pathway terms were driven by genes encoding MHC class II molecules (**Supp.** Fig. 2A-F). Within CD14+ monocytes, there was also enrichment of the matrisome-associated pathway, including increased amphiregulin (*AREG*) gene expression **(Fig. 3E, Supp.Table 8)**. We and others have previously reported AREG protein up-regulation in plasma in severe disease^14,15^. The present study suggests that CD14+ monocytes may contribute to this. The Reactome “Neutrophil Degranulation” module was also enriched in CD14+ monocytes in severe disease **(Supp.Table 8)**. *PLAC8* is a leading-edge gene in this pathway. In our dataset, it is significantly more highly expressed in severe disease **(Fig. 3E, Supp.Table 7)**. *PLAC8* over-expression makes cells, including immune cells, permissive for SARS-CoV-2 infection, and thus high expression of this molecule in patients with severe/critical disease may predispose them to worse outcomes^16^.

We also observed significant transcriptomic differences in NK cells between mild/moderate and severe/critical COVID-19. This included differential expression of genes relating to both TLR4 and TLR9 signalling pathways as well as those in the ‘Oncostatin M Pathway’ **(Supp.Tables 7-8)**. We previously reported upregulation of plasma protein levels of Oncostatin M in severe COVID-19^14^. This cytokine is known to regulate IL-6 and GM-CSF production, which have been previously implicated as drivers of severe COVID-19^17^. In dendritic cells, the sole pathway significantly associated with severity was the ‘Orexin Receptor Pathway’ which displayed a negative enrichment score in severe disease **(Supp.Table 8)**. This pathway was not significantly associated with severity in any other cell type, in contrast to the COVID-19 positive versus COVID-19 negative ESKD patient analysis, where we observed significant enrichment of this patient across multiple cell types.

In summary, mild/moderate and severe/critical disease were distinguished by transcriptional changes in numerous cell subsets. These were dominated by signals from B cell and monocyte subsets with a minor contribution from NK cells. By contrast, transcriptional changes in conventional alpha beta T cells were less able to distinguish mild/moderate and severe/critical disease.

### Temporal gene expression trajectories vary according to disease severity

The host response to infection is a dynamic process involving both the innate and adaptive immune systems. To understand these temporal dynamics in COVID-19, we performed longitudinal analysis of our multi-omic data. Cell type composition analysis revealed that most cell subtypes displaying an interferon-stimulated gene expression signature were significantly increased within the first week following symptom onset, and then gradually reduced over time (**Fig. 4A**). We observed significant increases in the relative abundance of some cell types persisting into weeks 2 and 3 following symptom onset (switched memory B cells, CD14 and CD16 monocytes, DC3, NK and CD8+ CTLs). I B cells, NK cells, CD4+ helper T cells, I CD8+ T cells, cytotoxic CD8+ T cells and Tregs were enriched in recovery samples. More generally, there was an increase in the relative abundance of adaptive immune cells over the course of the infection. As expected from a viral airway infection, compositions of antibody secreting B cells, predominantly of class-switched (IgG and IgA) antibody isotypes, were increased already in the first week after onset of disease, persisting for up to three weeks. Relative numbers of most CD4+ and CD8+ T cell subsets gradually decreased over time, with the exception of cycling CD4+ T cells (**Fig. 4A**).

**Fig. 4:**
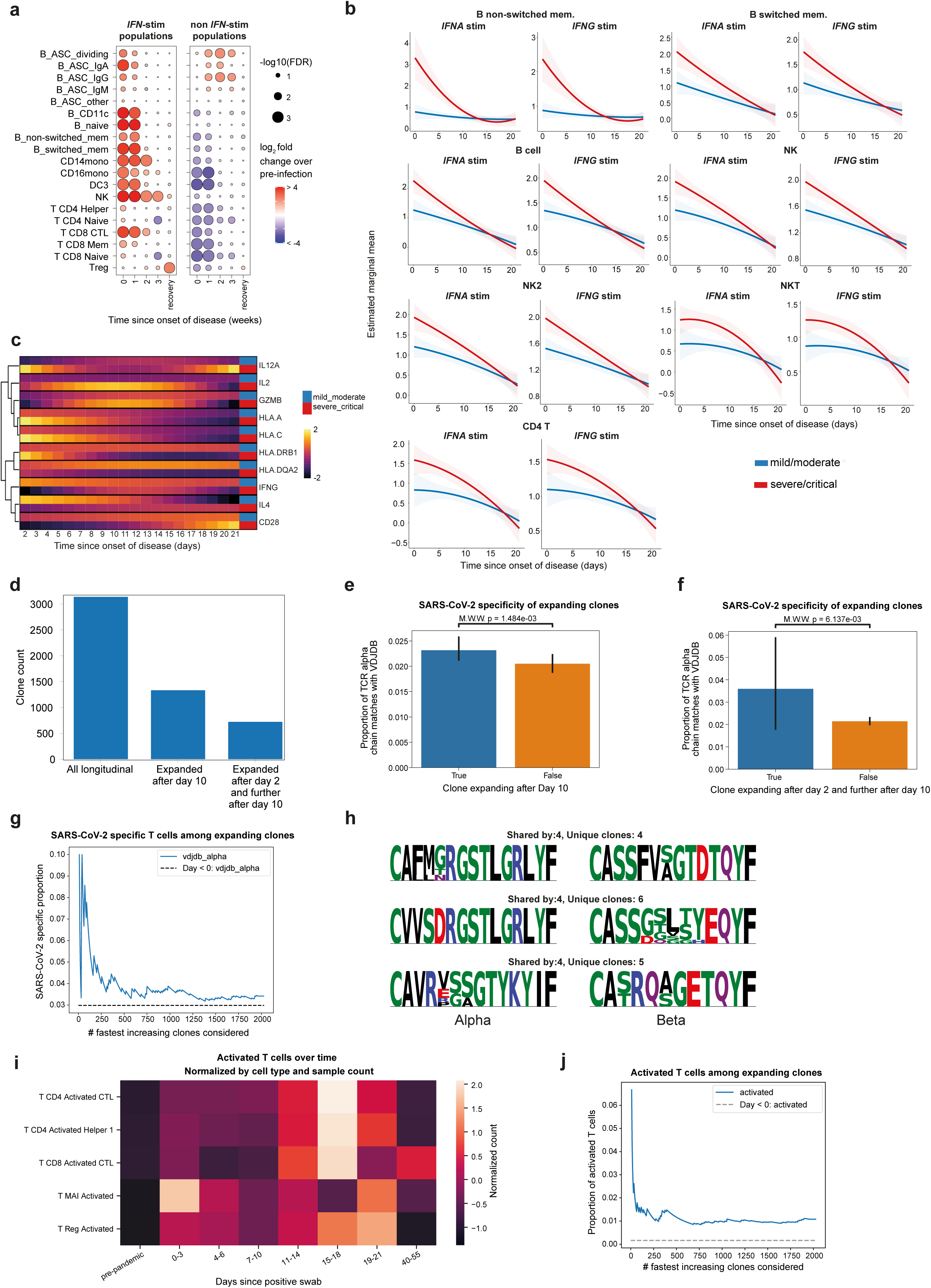
Longitudinal gene expression and TCR trajectories. **A)** Dot plots displaying the significant cell type abundance changes across COVID-19 infection compared to pre-infection samples for cells that have an interferon and non-interferon stimulated counterpart. Time since onset of disease is either time since display of first symptom or positive test (whichever is earliest). **B)** Estimated marginal mean of the effect of time from infection by severity group for the expression of IFN alpha and IFN gamma pathway genes for cell types with significant time x severity interaction. Time since onset of disease is either time since display of first symptom or positive test (whichever is earliest). **C)** Heatmap displaying 10 genes from multiple pathways (Allograft rejection, Asthma, Graft versus host disease, Type 1 diabetes, and Systemic Lupus Erythematosus) that had a significantly different temporal profile in mild vs severe disease (linear mixed model, FDR &lt; 0.05) in CD14 monocytes. Colour indicates LMM estimated marginal means over time, stratified by patient group (*n* = 130 samples from 37 individuals). Genes are clustered based on the temporal profile of the discordance between mild/moderate and severe/critical disease. Time since onset of disease is either time since display of first symptom or positive test (whichever is earliest). **D)** Absolute numbers of clones considered for longitudinal analysis and expanded clone counts. **E)** Proportion of SARS-CoV-2 specific clones among all clones, stratified by whether the clone expanded after day 10 following positive PCR test. Specificity was determined as a perfect match with a TCR alpha chain from the SARS-CoV-2 database VDJDB. Significance with two-sided Mann-Whitney test: p=0.0014. **F)** As for (C) but stratifying by whether a clone was expanded after day 2 and further after day 10. **G)** SARS-CoV-2 specific clone proportion among fastest increasing clones. Clones were sorted by decreasing expansion magnitude pre/post day 10 following positive PCR test (*Methods*). Baseline of matches with database from pre-pandemic samples shown in dashed line. Specificity was determined as a perfect match with a TCR alpha chain from the SARS-CoV-2 database VDJDB. **H)** Sequence logos of 3 most shared paired-chain TCR motifs, with number of individuals and number of unique clones sharing the motif mentioned. Letter height indicates frequency of AA at that position across T cells pertaining to the motif. AAs are coloured by side chain chemistry: Acidic (red), basic (blue), hydrophobic (black), neutral (purple), polar (green). AA: amino acid. **I)** Distribution of predicted activated T cells across days since positive swab result. T cell counts were normalized by number of cells and samples, cell states were predicted using Celltypist (*Methods*). **J)** Activated T cell state proportion among fastest increasing clones. Clones were sorted by decreasing expansion magnitude pre/post day 10 following positive PCR test (*Methods*). Baseline proportion of activated T cells from pre-pandemic samples shown in dashed line.

We next assessed the temporal patterns of gene expression changes during COVID-19 in ESKD patients and how these vary according to overall clinical course (defined by peak illness severity, binarised as mild/moderate or severe/critical). To achieve this, we performed longitudinal modelling using a linear mixed model with a time x peak severity interaction term. To reduce dimensionality, we analysed genes grouped together as modules according to pathway terms, using the Hallmark, Reactome and KEGG databases. A pathway with a significant time x severity interaction indicates that the pathway has a different temporal profile in mild/moderate versus severe/critical COVID-19. Our analysis revealed 183 pathways with significant (FDR <0.05) time x severity interactions (**Supp.Table 9**). Notably, the majority of the significant time x severity interactions were in B cells, accounting for 143 of the 183 significant pathways. Pathways in B cells and monocytes dominate those showing the 20 most significant time x severity interactions. The two pathways showing the most significant time x severity interaction were the interferon alpha and interferon gamma response in non-class switched memory B cells. Significant time x severity interactions for these pathways are also seen in B cells, class switched B memory cells, cytotoxic CD4+ T cells, NK cells, NK2 cells and NKT cells (**Supp.Table 9**). These results reflected quantitative differences in the temporal gradient of the interferon pathway response, with more severe COVID-19 disease showing higher interferon pathway response early in disease and a steeper decline over time (**Fig. 4B**).

In both CD14+ monocytes and B cells we found significant time x severity interactions for “allograft rejection” pathways’, and in CD14 monocytes for “graft versus host disease”, “asthma”, “type 1 diabetes” and “systemic lupus erythematosus”. Examination of the genes that make up these pathways revealed that these signals were largely driven by distinct temporal patterns of *HLA* expression in patients. In patients with a severe/critical clinical course, we observed steep downregulation of *HLA* class II gene expression over time, compared to either a relatively flat or mild upregulation in patients with a more benign course. *HLA* class I gene expression was higher in early disease in patients with severe/critical disease than in mild disease but fell further in late disease (**Fig. 4C and Supp.** Fig. 3A). Among other pathways that showed significant time x severity interactions, we noted the “KRAS signalling up” and “MYC targets” in B cells only, likely reflecting time dependent changes in their proliferation during infection that vary according to severity (**Supp.Table 9**).

These results illuminate how modelling the temporal component provides additional insights by identifying time-dependent severity associations with gene expression that are not apparent in single time-point cross-sectional analyses. Thus, transcriptomic changes are dependent both on time and severity, and the interplay of two, underscoring the importance of serial sampling in gaining a complete picture of the host immune response in COVID-19.

### Longitudinal TCR dynamics

Given the central role of T cells in antiviral adaptive immunity, we next evaluated clonal T cell dynamics of SARS-CoV-2 infection. The longitudinal nature of our study and single-cell resolution enabled us to be specific in determining paired-chain clones that expanded over the course of COVID-19. A total of 3,137 unique TCR clones that appeared in two or more serial samples from the same patient were used to quantify clonal expansion. To increase the probability of identifying TCR clones specific to SARS-CoV-2, we focused on clones that were not present in pre-infection samples, thereby limiting the presence of cross-reactive or bystander T cells. We found that 42% of clones sampled longitudinally had increased clonal frequency following day 10 after a positive swab, and that 23% showed a marked expansion where they increased after day 2 of the positive swab, and further after day 10 (**Fig. 4D**, **Supp.** Fig. 3B-C, *Methods*). To investigate whether these clonal expansions were directed against SARS-CoV-2, we cross-referenced SARS-CoV-2 specific TCR sequences from the VDJDB database^18^ and measured the overlap with clones identified in more than one serial sample within an individual. Clones expanding after day 10 were significantly enriched in SARS-CoV- 2 specific TCR alpha chains (p=0.0014, two-sided Mann-Whitney test, **Fig. 4E**) compared to their non-expanding counterparts, while those fulfilling the stricter dual criteria above had an almost two-fold increase in antigen-specific TCRs (p=0.006137, **Fig. 4F**). We next tested the relationship between magnitude of expansion of the longitudinally identified clones and SARS-CoV-2 specificity. The fastest expanding clones had the highest proportion of SARS-CoV-2 specific TCR alpha chains (**Fig. 4G**), indicating that we are capturing the adaptive immune response to COVID-19. This SARS-CoV-2 specificity estimate is likely a lower bound to the true number, as experimental data from the database is based on assays with many fewer SARS-CoV-2 peptides than the number of naturally occurring viral antigens. Thus, of the expanding sequences that we recovered that do not match the database, many more are likely to be virus specific.

Further leveraging our single cell data, we looked for patterns in the TCRs of expanding clones that might be shared across donors. We applied the tool Cell2TCR^19^ to our expanded clones (excluding MAIT cells, *Methods*) and found 99 public TCR motifs, where a public TCR motif denotes a group of clonotypes with sufficient sequence similarity to likely recognise the same epitope that was found in two or more patients. Moreover, six TCR motifs were shared between three donors and three motifs between four donors, a scenario which is highly unlikely for randomly sampled TCR clones and provides evidence of strong selective pressure on the adaptive immune response to a common pathogen (**Fig. 4H**). As we had recruited patients during two distinct phases of the pandemic, we hypothesised that certain TCR motifs might be specific to a particular viral strain and exhibit sharing only across donors from the same cohort (e.g. sampled in 2020 or 2021, respectively). Of the 99 public TCR motifs, 64% had donors from a single cohort, including two motifs of **Fig. 4I** with four donors each. Furthermore, 18% of public motifs contained at least one SARS-CoV-2 specific TCR sequence, underscoring the sensitivity of this approach to analyse the antigen-specific response. Our findings are in strong agreement with evidence from a recent SARS-CoV-2 Human Challenge Study^19^, involving deliberate infection of healthy individuals with SARS-CoV-2, which had shown that the antigen-specific response included convergent paired-chain immune receptor motifs. We thus replicate the challenge study results in the context of natural infection and in a larger cohort comprising a clinically relevant vulnerable group consisting of older individuals with underlying comorbidities, which included cases of severe/critical COVID- 19.

As our data was longitudinal, we next investigated the presence of time-restricted, activated T cell types characterised in the context of COVID-19 by Lindeboom *et al*^19^. Activated T cell states were found to be indicative of *de novo* T cell activation and harbouring SARS-CoV-2 specific TCR sequences. Application of the automated cell state annotation tool Celltypist^9^ revealed 1,927 activated T cells, spanning the CD4+, CD8+, regulatory and MAIT cell compartments and found among 58 ESKD COVID-19 patients (**Supp.** Fig. 3D). When normalising the counts by cell type and sample numbers and aggregating across time points, we observed a striking lack of activated T cells in pre-pandemic as well as convalescent COVID-19 samples (**Fig. 4H**). While MAIT cells and regulatory T cells showed a relative enrichment during the first week after positive PCR test, most activated CD4+ and CD8+ appeared only after 10 days. All activated T cell types remained detectable three weeks after positive PCR test but had mostly disappeared again by the time convalescent samples were taken, highlighting the transient nature of these cell states. Activated T cells were further over- represented among the most expanded clones (**Fig. 4J**). This is in line with results from the clinical trial in Lindeboom *et al*, where activated MAIT cells could be detected as early as 3 days after exposure to the virus, and circulating activated T cell abundance peaked 10 -14 days after exposure to the virus, with return to baseline after 28 days^19^.

### Corticosteroids induce dexamethasone-related monocytes in COVID-19

By the time of the 2021 COVID-19 wave in the UK, glucocorticoid administration with dexamethasone had become standard clinical practice following randomised clinical trials demonstrating that it reduced mortality in patients with COVID-19 requiring supplemental oxygen^3^. Corticosteroids are known for their broad immunosuppressive effects through several different mechanisms, including inhibiting the release of proinflammatory cytokines^20^. *In vitro* experiments have suggested that monocytes and macrophages treated with glucocorticoids can exhibit both anti-inflammatory and inflammation-resolving properties^21^. The effect of corticosteroids on human immune responses at single cell level *in vivo* has not been studied. Of the 16 patients in the 2021 Cohort, 7 received steroid treatment **(Supp.Table 1).** Patients receiving steroids all had a peak illness severity of severe or critical. This provided us with an opportunity to investigate the effects of steroids at the single cell transcriptomic level over the course of their treatment.

Whilst exploring the innate immune response, we isolated the monocyte compartment from the rest of the data and sub-clustered these cells. We identified the emergence of a subset of monocytes that were only seen in COVID-19 positive, severe/critical cases but not in patients with mild/moderate disease (**Fig. 5A-B**). These cells were clearly demarcated on UMAP plots as a distinct population. Differential gene expression analysis between all subsets of monocytes showed this population had transcriptional similarities with monocytes treated *ex vivo* with dexamethasone^22^ (**Fig. 5C**). Compared to classical CD14 monocytes and IFN stimulated CD14 monocytes, the dexamethasone-related monocytes (Dex. mono) had lower expression of markers of inflammation such as *JUN* and *CXCL8* as well as lower expression of antigen presenting markers *HLA-DRA* and *HLA-DRB5*. Conversely, they showed higher expression of genes relating to anti-inflammatory actions (*CD163* and *ADAMTS2)*, anti- oxidation (*SLC1A3* and *SESN1),* migration (*FPR1* and *MTSS1*) and phagocytosis (*MFGE8* and *MRC1)* (**Fig. 5C**). Notably, these cells were only present in patients recruited in the 2021 Cohort, suggesting they were a direct effect of glucocorticoid treatment and not a consequence of severe COVID-19 itself (**Fig. 5A**).

**Fig. 5:**
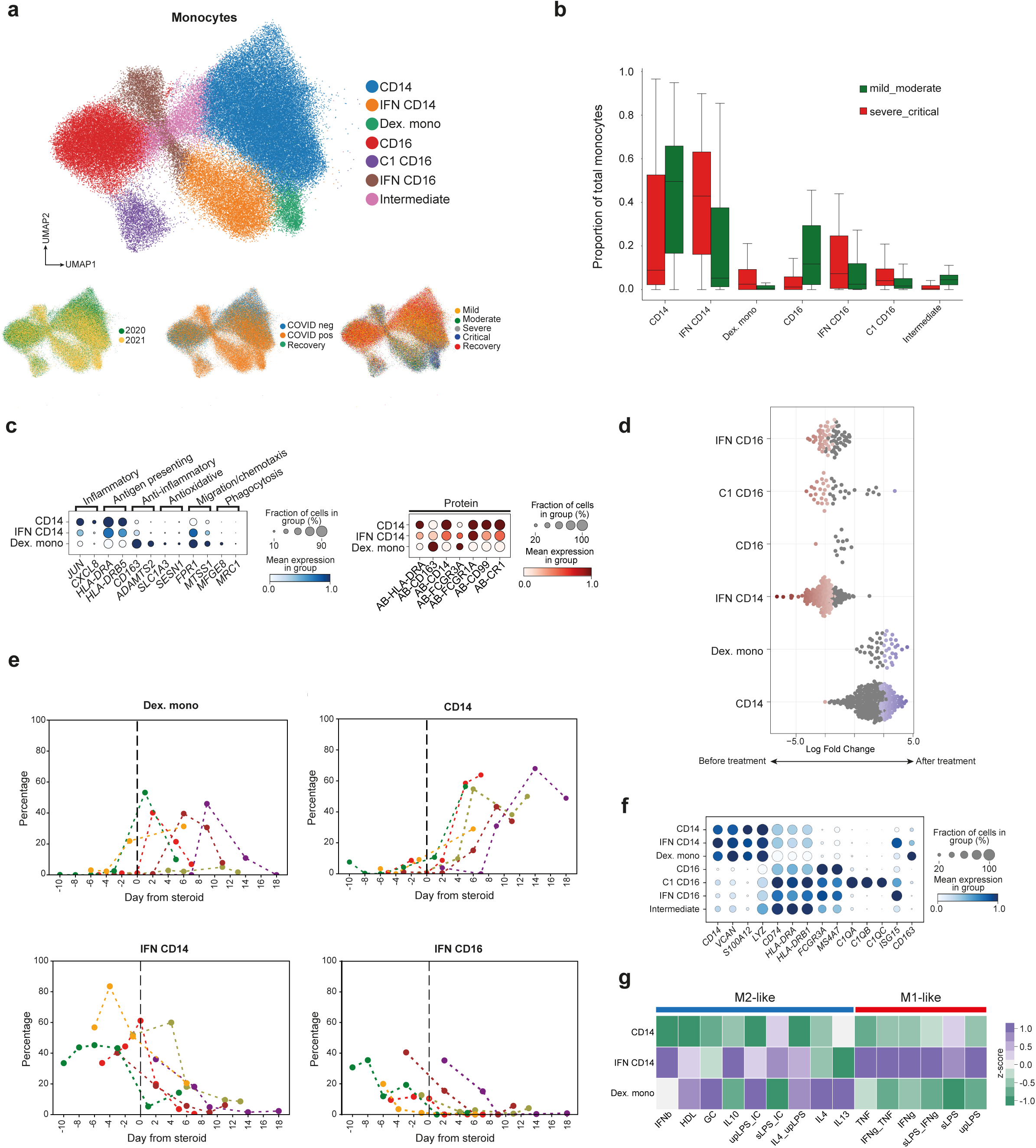
**Dexamethasone treatment promotes steroid associated monocytes**. **A)** UMAPs displaying all subsets of monocytes; coloured by subset (top), patient cohort (bottom left), COVID-19 status (bottom middle) and severity (bottom right). **B)** Bar charts displaying proportions of all monocytes grouped by mild/moderate and severe/critical severity. **C)** Dot plots displaying gene expression (left) and protein expression (right) in CD14 monocytes, IFN-stimulated CD14 monocytes and the dexamethasone associated monocytes (Dex. mono). **D)** Beeswarm plot displaying the differential abundance of monocyte subsets for samples from patients that were administered steroids, before and after treatment. **E)** Line charts displaying the percentage of monocyte subsets across the days before and after administration of steroids. Line colours represent different patients. **F)** Dot plot displaying the expression of monocyte marker genes in all monocyte subsets. **G)** Heat map displaying gene module scores for CD14 monocytes, IFN-stimulated CD14 monocytes and the dexamethasone associated monocytes (Dex. mono).

We formally tested the effect of glucocorticoids on differential cell abundance across the monocyte clusters using MiloR, accounting for time from infection^23^. We noted that both CD14 monocytes and the dex. monos were significantly enriched after glucocorticoid treatment, and the IFN-stimulated CD16 monocytes, C1 CD16 monocytes and IFN-stimulated CD14 monocytes were significantly enriched before treatment (**Fig. 5D**). Using the longitudinal data from only the individuals who were given glucocorticoid treatment, we evaluated the percentage of different monocyte subsets prior to and in the days after treatment. We found that, after glucocorticoid administration, there was a trend towards an increased relative abundance of the dex. monos and CD14 monocytes, whilst there was a decrease in both IFN-stimulated monocyte populations (**Fig. 5E**). No trends were observed in other cell types (**Supp.** Fig. 4 and 5). These results suggest that, along with promoting the emergence of the dex. monos, glucocorticoids could facilitate a reduction of the abundance of interferon-stimulated monocytes in severe COVID-19 infection.

The dex. monos displayed high RNA and protein expression of CD163 (**Fig. 5C and 5F**) a scavenger receptor that is frequently used to mark ‘alternatively activated’ or ‘M2’-like macrophages^24^. These macrophages possess regulatory functions which can suppress immune responses and reduce inflammation^25^. Macrophages treated with glucocorticoids have been shown to drive the polarisation of macrophages towards an ‘alternatively activated’/M2-like phenotype^26^. These findings prompted us to further assess transcriptional programs of the dex. monos. We performed pathway enrichment analysis on all monocytes based on 15 different macrophage stimulation signatures^27^. The dex. monos were more associated with transcriptomic patterns associated with stimulation with IL-13, IL-4, ultra-pure LPS+immune complex, glucocorticoid stimulation, supporting their similarity to M2-like macrophages (**Fig. 5G**). The temporal emergence of the dex. monos and their presence only in the 2021 Cohort strongly suggest that the changes we observed were driven by dexamethasone treatment rather than disease severity.

## Discussion

Here, we performed CITE-seq and immunoreceptor profiling in ESKD patients with COVID-19 to longitudinally profile the circulating immune cell changes associated with COVID-19 in two temporally distinct cohorts. A unique aspect of our study was the 2021 cohort, where we obtained longitudinal PBMC samples from patients with COVID-19 who were originally sampled as COVID-19 negative controls during 2020 but subsequently became infected during 2021. As a result, we were able to perform intra-individual analysis of the host immune cell PBMC transcriptome comparing pre-infection with acute infection, thus minimising the impact of confounding factors. Another strength of our study was the inclusion of patients of diverse ancestries.

Using a multi-omic approach, we identified COVID-19-associated changes in the cellular composition of PBMC in ESKD patients including increased relative abundance of naive B cells and a decreased relative abundance of total monocytes, CD8+ memory and CD4+ CTL T cells. This decrease in the relative numbers of circulating monocytes following infection was also observed in a recent experimental medicine challenge study, involving deliberate infection of healthy individuals with SARS-CoV-2^19^. COVID-19 was associated with widespread transcriptomic changes in a wide variety of cell types. Many of these reflect the activation of inflammatory pathways, including the type 1 interferon pathway and cellular activation and proliferation. Overall, the COVID-19-associated changes in ESKD patients were similar to those reported in other studies, but we did identify some changes that, to our knowledge, have not been previously reported. For example, gene expression pathway analysis highlighted significant negative enrichment of the “Orexin Receptor Pathway’’ in COVID-19 positive ESKD patients versus uninfected ESKD patients across several innate immune cell types. Many of the leading-edge genes contributing to the “Orexin receptor pathway” term are also involved in other pathways, which makes interpretation of this finding more challenging. Orexin receptor signalling is well-characterised in neurological diseases such as narcolepsy, but there is also evidence that Orexins can have immunological effects that may be relevant in the context of COVID-19^28,29^.

We also identified numerous pathways associated with COVID-19 severity in ESKD, particularly in monocytes and B cells. Notably, we found many more pathways associated with severity in B cells than in T cells. In addition, severe COVID-19 was associated with a higher relative abundance of antibody-secreting B cells and with higher expression of genes involved in cell division. In CD14+CD16- monocytes, we observed elevated *PLAC8* expression in severe COVID-19. High *PLAC8* expression makes lung cells more permissive for SARS-CoV- 2 infection *in vitro*^16^. In line with this, a genome-wide CRISPR knockout screen identified PLAC8 as an essential factor for infection with a different coronavirus, swine acute diarrhoea syndrome coronavirus (SADS-CoV)^30^. While SARS-CoV-2 predominantly infects epithelial cells, it has also been detected in macrophages and T cells^31^. Together with this work on the cell biology of *PLAC8* in viral infection, our observation that expression changes with disease severity in monocytes raises the possibility that modulating *PLAC8* expression may provide a therapeutic opportunity to prevent infection of both epithelial and immune cells.

Multi-omic measurements allowed us to identify instances of negative correlation between immune cell gene expression and levels of the corresponding plasma protein. For example, in severe COVID-19 the most down-regulated gene in monocytes was *TNF* (encoding TNF-alpha), yet conversely, TNF-alpha was significantly upregulated in the plasma from the same blood draw. Potential explanations for this uncoupling include negative feedback, or that other cell types could be contributing to circulating TNF-alpha pool (e.g. endothelial cells, tissue macrophages). This observation underlines the complementary value of combining multi-omic data, since plasma proteins reflect protein production by a wide variety of tissues other than blood cells^32^. Since our data are observational, we cannot determine if elevated circulating TNF-alpha is a cause or a consequence of severe COVID-19. Nevertheless, since TNF-alpha blocking drugs are in routine clinical use in inflammatory diseases such as rheumatoid arthritis, our results suggest there may be value in evaluating the repurposing of these medications for COVID-19^33^.

Longitudinal analysis of changes in cell type abundance showed a peak in cells showing an interferon-activated gene signature in the first week of illness followed by a waning, again consistent with that seen in other work^19^. Gene expression pathways which displayed distinct temporal profiles according to clinical severity were predominantly found in B cells and monocytes.

Longitudinal analysis also revealed the time-restricted appearance and expansion of T cells with likely SARS-CoV-2 specificity. Leveraging the large number of longitudinal samples, we identified the emergence of public T cell clones with a restricted TCR repertoire that are shared across patients. Cross-referencing the TCRs with SARS-CoV-2 specific databases, we found most matches among T cells that strongly expand in the days following a positive PCR test, and a significant enrichment of matches compared to pre-pandemic samples. Our results support the hypothesis that both TCR chains together determine antigen specificity, which we were able to capture jointly using single-cell sequencing, as expanding clones could further be grouped into shared TCR motifs with high sequence similarity in both chains. In addition, a recently activated T cell phenotype is overrepresented in that same population. We expand on existing knowledge from the first human COVID-19 challenge study by analysing a larger patient cohort, which included cases of severe disease, as well as replicating several key findings in the context of natural infection. In addition, we identified longitudinal expansion of some clones that are not recorded as SARS-Cov2-specific in the VDJDB database. These could potentially be previously undescribed SARS-CoV2-specific T cells, given that we analysed a large number of longitudinal samples that included patients with severe/critical disease and also diverse ancestry. An alternative explanation is that the expansion of some TCR clones was driven by bystander activation secondary to the inflammatory milieu. Thus, our study generates hypotheses for further functional work.

An important finding of this study was the identification of a distinct population of monocytes that emerged after glucocorticoid therapy. These were not observed in any patients in the 2020 Cohort, before the introduction of glucocorticoid therapy as standard of care, and their emergence in the 2021 Cohort occurred rapidly after glucocorticoid administration. These findings could have implications beyond COVID-19. Corticosteroids are frequently used to suppress inflammation and they have pleiotropic effects on immunity that are not fully understood despite their long-standing clinical use. Therefore, being able to probe the effects of steroid treatment in humans using single cell resolution multi-omic technologies provides potential insights into their mode of action. This may facilitate rational drug design of compounds that target these pathways without steroid side effects. Here, we demonstrated that glucocorticoids could promote the emergence of a transcriptionally distinct subpopulation of monocytes. An outstanding question remains the functional properties of these steroid-induced cells such as their ability to traffic to tissues and modulate inflammatory responses.

ESKD patients have increased susceptibility to sepsis and impaired vaccination response (for example, hepatitis B vaccination)^8,34^. Reports on neutralising activity against the delta variant following SARS-Cov-2 vaccination showed an impaired response of haemodialysis patients compared with healthy controls in response to the AZD1222 vaccine^35^ and an impaired neutralising response to the omicron variant with heterologous boost regimes consisting of two doses of the AZD1222 followed by the mRNA vaccine BNT162b2^36^. Vaccination strategies have been very successful in reducing morbidity and mortality, but widespread transmission of SARS-CoV-2 continues. As new variants emerge, certain patient groups, such as those with ESKD will have a higher risk of both contracting infection and of experiencing a severe disease course, underscoring the importance of studying such patient groups.

Our study has some limitations. Our data are observational and thus cannot delineate whether changes in cell populations or gene expression are pathogenic drivers or downstream consequences of the systemic inflammatory response. Observational data are also vulnerable to the effects of confounding factors. Our use of paired pre-infection and infection samples in the analysis of the Wave 2 partially mitigates this, but unknowable confounders such as viral exposure at the time of infection may nevertheless impact the magnitude of the host immune response. In addition, we studied peripheral blood immune cells due to accessibility, but these may not always reflect those at the site of tissue inflammation. We did not have a comparator group of ESKD patients with another infection, so we cannot determine whether the changes we observed are specific to COVID-19. Finally, this was a single centre study.

In summary, we characterised the longitudinal host immune response in COVID-19 in a clinically vulnerable group through multi-omic technologies. These data illuminate the temporal dynamics of the response to infection, and how these diverge in mild versus severe disease. Our results reveal the impact of glucocorticoid therapy, with the emergence of a specific monocyte subpopulation following treatment. The data here will provide a valuable resource for the research community.

## Data Availability

Data will be available online at https://covid19cellatlas.org/

## Acknowledgements

This work was funded by a UKRI-DHSC COVID-19 Rapid Response Rolling Call (MR/V027638/1) (to J.E.P.), funding from the UK Coronavirus Immunology Consortium (UK-CIC) and Wellcome Human Cell Atlas Strategic Science Support (WT211276/Z/18/Z), and the NIHR Imperial Biomedical Research Centre (BRC). The views expressed are those of the authors and not necessarily those of the NIHR or the Department of Health and Social Care. M.H. is funded by Wellcome (221052/Z/20/Z, 215116/Z/18/Z), the Lister Institute for Preventive Medicine, and NIHR and Newcastle Biomedical Research Centre. L.M.D is supported by the European Union’s Horizon 2020 research and innovation programme under the Marie Skłodowska-Curie grant agreement No 955321. J.E.P. is supported by a fellowship from the Medical Research Foundation (MRF-057-0003-RG-PETE-C0799). M.C.P. is a Wellcome Trust Senior Fellow in Clinical Science (212252/Z/18/Z). C.L.C was supported by an Auchi Clinical Research Fellowship.

## Author contributions

Conceptualisation, M.B., S.A.T, M.H, M.R.C, J.E.P; Investigation, E.St., N.B.B, M.C, A.P., E.P., T.M and A.P. ; Methodology, J.R.F, B.J.S., J.G., Resources, C.L.C., N.M-T., M.P. S.M., M.W. and E.Sa. ; Formal analysis, E.St., E.M-D., L.M.D., R.G.H.L., Z.K.T., W.M.T., S.B. and D.C.T.; Writing, E.St., E.M-D., L.M.D., R.G.H.L., Z.K.T., W.M.T., D.C.T. and J.E.P.; Editing: M.C.P., M.B., S.A.T., M.H. and M.R.C.; Supervision, M.B., S.A.T., M.H., M.R.C, D.C.T. and J.E.P.

## Supplemental information

Figures S1-S6 and Tables S1-S9.

## Materials and Methods Ethical approval

All participants (patients and controls) were recruited from the Imperial College Healthcare NHS Trust Renal and Transplant Centre and its satellite dialysis units, London, United Kingdom, and provided written informed consent prior to participation. Study ethics were reviewed by the UK National Health Service (NHS) Health Research Authority (HRA) and Health and Care Research Wales (HCRW) Research Ethics Committee (reference 20/WA/0123: The impact of COVID-19 on patients with renal disease and immunosuppressed patients). Ethical approval was given.

## Patient cohorts

We recruited two cohorts of ESKD patients with COVID-19. All patients were on haemodialysis prior to acquiring COVID-19. The first cohort (‘2020/Wave 1’) were recruited during the initial phase of the COVID-19 pandemic (April-May 2020). We collected 61 serial blood samples during acute infection for 21 patients with COVID-19. Three samples were collected for 19 of these patients; two samples were collected for the other two individuals. We also contemporaneously recruited 37 non-infected ESKD patients on haemodialysis to provide a control group.

The second cohort (‘2021/Wave 2’) were recruited during the resurgence of cases in January-March 2021. This cohort, which consisted of 16 ESKD patients with COVID-19, had all been recruited as part of the COVID-19 negative control group during the 2020 wave, and so a pre-infection sample collected in April/May 2020 (8-9 months preceding infection) was also available for 13 patients. For these patients, samples were systematically acquired at regular intervals (median 5 samples per patient, collected every 2-3 days over the course of the acute infection). Additionally, for 10 of these 16 patients, we acquired convalescent samples approximately 2 months following the acute COVID-19 episode. 3 individuals in this cohort had received one dose of the COVID-19 vaccine, however their first blood sample was taken within an average of 5 days so there was an unlikely chance this had an effect on their immune response to the infection.

## Clinical severity scores

Severity scoring was performed based on WHO classifications (WHO clinical management of COVID-19: Interim guidance 27 May 2020) adapted for clinical data available from electronic medical records. ‘Mild’ was defined as COVID-19 symptoms but no evidence of pneumonia and no hypoxia. ‘Moderate’ was defined as symptoms of pneumonia or hypoxia with oxygen saturation (SaO2) greater than 92% on air, or an oxygen requirement no greater than 4 L/min. ‘Severe’ was defined as SaO2 less than 92% on air, or respiratory rate more than 30 per minute, or oxygen requirement more than 4 L/min. ‘Critical’ was defined as organ dysfunction or shock or need for high dependency or intensive care support (i.e. the need for non-invasive ventilation or intubation). Severity scores were charted throughout a patient’s illness. We defined the overall severity/clinical course for each patient as the peak severity score that occurred during the patient’s illness.

## PBMC isolation protocol

Peripheral blood mononuclear cells (PBMCs) were obtained by density gradient centrifugation using Lymphoprep (STEMCELL Technologies, Canada). Approximately 20 ml of blood were diluted 1× with phosphate buffered saline (PBS) with addition of 2% fetal bovine serum (FBS) and layered on top of 15 ml of Lymphoprep solution. The samples were then centrifuged at 800 g for 20 min at room temperature without break. PBMCs were collected from the interface and washed twice with PBS/2%FBS. PBMCs were cryopreserved in 1 ml freezing medium (FBS 10% DMSO) and stored in liquid nitrogen.

## PBMC processing and CITEseq

### Samples collected during 2020 wave

Frozen PBMCs were thawed by adding a small volume of ice-cold PBS to PBMC samples and transferred to a falcon tube containing 35 mL of ice-cold PBS. Samples were then centrifuged and counted. Dead cells were removed using the EasySep Dead Cell Removal kit (Stem Cell Technologies) according to the manufacturer’s protocol. Cells were then counted again and 40,000 cells from each sample were pooled together in batches of seven with the aim for each pool to contain ∼300,000 cells, ensuring each pool had a different combination of genotypes for simple demultiplexing. Pooled cells were then stained with Fc Receptor Blocking Solution (Biolegend) and then with TotalSeq™-C Human Universal Cocktail V1.0 (Biolegend) according to the manufacturer. Cells were then washed once with PBS and then counted. Each pool was loaded across two channels of a Chromium Chip (10x Genomics), using Single Cell 5’ V2 kits, to achieve a recovery of 10,000 cells per sample.

### Samples collected during 2021 wave

Frozen PBMCs were thawed at 37°C until a small ice crystal remained. Samples were then transferred to another tube and ten times the volume of pre-warmed RF-10 media (RPMI (Sigma) supplemented with 10% (v/v) fetal calf serum (Life technologies), 100U/ml Penicillin (Sigma), 100 µg/ml Streptomycin (Sigma) and 1% (v/v) L-Glutamine) was added dropwise. Cells were then centrifuged and counted. Dead cells were removed using the EasySep Dead Cell Removal kit (Stem Cell Technologies) according to the manufacturer’s protocol. Cells were then counted again and 250,000 cells from each sample were pooled together in batches of four using a leave-one-out strategy for simple demultiplexing. Pooled cells were then stained with Fc Receptor Blocking Solution (Biolegend) and then with TotalSeq™-C Human Universal Cocktail V1.0 (Biolegend) according to the manufacturer. Cells were then washed three times with Flow Buffer (Dulbecco’s phosphate buffered saline (PBS)(Sigma) supplemented with 2% (v/v) FCS and 2mM EDTA (Sigma)) and then counted. Each pool was loaded across two channels of a Chromium Chip (10x Genomics), using Single Cell 5’ V2 kits, to achieve a recovery of 10,000 cells per sample.

### Library Preparation and Sequencing

Gene expression, cell surface protein, TCR and BCR libraries were generated according to the manufacturer’s protocols. All libraries were sequenced using a NovaSeq 6000 to achieve a minimum of 20,000 reads per cell for gene expression libraries and 5,000 reads for cell surface protein, TCR and BCR libraries.

### Bioinformatics Pre-processing

We jointly aligned the antibody-derived tags (ADT) and gene expression libraries from CITE-seq experiments using *CellRanger 4.0*, using the reference 10X Genomics provided with the release of *CellRanger 3.0*, and the ADT barcode reference provided by the supplier. Single cell TCR and BCR sequencing data was aligned using *CellRanger 4.0* using the GRCh38 VDJ reference provided by 10X Genomics. We used *Seurat* V4.1.0 ^37^ to import gene expression and ADT counts. Low quality cells were excluded by removing droplets with either fewer than 1000 RNA UMIs, or fewer than 200 RNA features detected, or with more than 10% of their RNA UMIs mapping to mitochondrial genes. *SoupX* ^38^ was used to remove signals from ambient RNA and background antibody staining. SoupX parameters ‘soupQuantile’ and ‘tfidfMin’ were set to 0.25 and 0.2, respectively, and lowered by decrements of 0.05 until the contamination fraction was calculated using the ‘autoEstCont’ function. Corrected gene expression and ADT counts were then scaled to 10000 UMIs per cell and log1p transformed.

### Sample demultiplexing

We used *souporcell v2.0* ^39^ to perform genotype-based demultiplexing of pooled PBMC libraries to assign donor identifiers to each single cell transcriptome. To ensure high reproducibility of the genotype-decomposition, we merged the sequencing data from each set of replicates of the same donor pool prior to *souporcell* analysis. We used *pysam v0.17.0* to amend cell barcodes with original library identifiers and to merge bam files. Using the merged bam files, we ran *souporcell* using the provided set of common variants, with remapping disabled and with the appropriate number of expected genotypes. To assign a donor identifier to each *souporcell* genotype cluster we leveraged the pooling strategy of donors per library which was designed in such a way that every donor was present in a unique combination of pools. We used the *cardelino* R package ^40^ to import genotypes and perform pairwise comparisons of all identified *souporcell* genotype clusters, to identify highly similar genotype clusters in different pools that likely originated from the same donor, which was then given a donor label based on the combination of pools in which the genotype was detected. Genotypes that were not resolvable due to missing or low-quality data, were excluded from downstream analyses.

We detected a total of 1337786 cells with at least 200 genes quantified. We next applied stringent filtering on cell quality to remove cells with more than 10% mitochondrial reads and cells with less than 1000 UMIs quantified. In addition, we only kept cells with a genotype / patient id assignment using souporcell, and that did not cluster in doublet enriched leiden clusters during the manual annotation process. This resulted in a dataset of 588389 high-quality cells from 63 patients and 198 samples that were used for the reported analyses.

### Single-cell Quality Control - Myeloid and non-immune haematopoietic cell compartment

Annotation of myeloid and progenitor compartment was performed using *scanpy* ^41^ (v1.8.2). The dataset was initially normalized, and log transformed, and then filtered for highly variable genes (*scanpy.pp.highly_variable_genes; min_mean=0.0125, max_mean=3, min_disp=0.5*) and scaled (*scanpy.pp.scale, max_value=10*). Dimensionality reduction was performed using principal component analysis (PCA; *scanpy.tl.pca*), and integration was done using *harmony* ^42^ (*harmonypy*, v0.0.6). Clustering was performed using the Leiden ^43^ algorithm (*leidenalg,* v0.8.9). The marker genes for each cluster were examined using the function ‘*scanpy.tl.rank_genes_groups*’ and each cluster was manually annotated.

### Single-cell Quality Control - T and NK cell compartment

The T and NK cell compartment quality control and annotation was performed using the *Seurat* (v4.1.1) workflow ^37^. The expression data was normalized, and log transformed (normalized to 10,000 counts per cell), 2000 highly variable genes were selected (*FindVariableFeatures* function, *selection.method = ‘vst’*), from which TCR and V(D)J genes were excluded. Prior to scaling the gene expression data, unwanted sources of variation in the form of total read count and percentage of mitochondrial genes were regressed out (using *ScaleData* function, *vars.to.regress* argument). Integration of sequencing samples (*‘orig.ident’*) using *harmony* ^42^ (v1.0) was carried out on the first 30 principal components of the expression data. K-nearest neighbors (KNN) and shared nearest neighbors (SNN) graphs were calculated from the harmony adjusted PCs (*FindNeighbours* function). Finally, cells were clustered using the Leiden algorithm (*FindClusters* function, *method=’igraph’, algorithm=4*, requiring *‘leidenalg’* python package) and visualized via non-linear dimension reduction UMAP. Clusters were manually annotated using canonical marker genes through an iterative process of re-clustering, annotation, identification of potential doublets (presence of distinct cell type marker genes) and re-clustering. CITE-seq marker proteins CD45RA and CD45RO were used in the annotation of T EMRA and other memory T cells respectively. All other markers used for annotations were based on mRNA expression data.

### Single-cell Quality Control - B cell compartment

The B cell compartment was integrated using *scVI* (v.0.19.0) ^44^ with sequencing samples (‘*orig.ident*’) as the batch key and raw count data as input. Samples from two individuals were observed to not integrate well and they were subsequently identified to be samples from patients with benign chronic lymphocytic leukaemia and were removed from all downstream analyses (**Table S1**). Percentage mitochondrial content and total counts were provided as continuous variables to the *scVI* model. Feature selection prior to setting up the scVI model was performed as per standard procedures in *scanpy.pp.highly_variable_genes* with *min_mean=0.0125, max_mean=3, min_disp=0.5*, using the log transformed normalized expression data (normalized to 10,000 counts per cell). BCR V(D)J genes were also removed from the highly variable features. Expression of canonical B cell and ASC marker genes and non-B cell markers were then assessed to manually determine potential multiplets, over iterative rounds of sub-clustering. The annotations were also assessed against a publicly available bulk RNAseq gene set of major PBMC cell types ^45^. In addition, the single-cell scores computed after enrichment of the bulk RNA-seq signatures were fitted into a two-component Gaussian mixture model (*max iter=1000, covariance_type=’full’*) which distinguished ASCs from non-ASC B cell clusters. Subsequent sub-clustering and annotations were performed on the ASCs and non-ASCs separately. To annotate the non-ASC cell clusters, mRNA and surface molecule expression for select targets (CITE-seq; CD11C and CD27), along with the Monaco et al. ^45^ peripheral blood B cell signatures. Isotype usage was checked using the single-cell and BCR-seq information and used to manually update the cell type annotations, ensuring that naive B cells, non-switched memory B cells and IgM ASCs are only associated with IgM and/or IgD while switched memory B cells and IgA/IgG ASCs are only associated with IgG/IgA isotypes. Other antibody isotype expressing ASCs (IgD/IgE) are labelled as ‘*B_ASC_others*’.

### Integration of Olink Plasma Proteomics

A subset of Wave 1/2020 Cohort (45 individuals, 85 samples) had plasma proteomics measures from 5 Olink Proteomics Target 96 panels: ‘cardiometabolic’, ‘cardiovascular 2’, ‘cardiovascular 3’, ‘inflammation’ and ‘immune response’. The Olink proteomics data for these samples has previously been described^14^.

### Longitudinal analysis

We defined time from infection as the time from first symptoms, or time from first positive nasal swab if the latter preceded symptoms (since some cases of COVID-19 were identified by screening procedures in place for patients attending haemodialysis).

For longitudinal analysis of enrichment of MSigDB (v7.5) Hallmark, KEGG and Reactome genesets ^46^, the single-cell data was separated to each cell type and the raw count data was aggregated by sample using ‘*scuttle::aggregrateAcrossCells*’ (v1.9.0). Only samples with more than 10 cells were used for downstream analysis. The pseudo-bulked data was then log transformed and normalized using ‘*scuttle::logNormCounts*’ and converted to a module score using ‘*Seurat::AddModuleScore*’. The module scores were then tested for differential enrichment over time according to severity strata, using a general linear mixed-effect model with ‘*lme4:lmer*’ using the following formula:

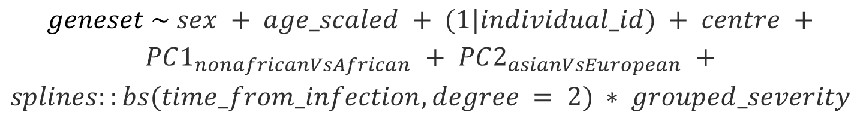

“Grouped severity” represents overall clinical course, defined by peak illness severity, binarised into either severe/critical or mild/moderate. The estimated marginal means for the first 21 days from infection for the relevant genesets were computed using ‘emmeans::emmeans’ with ‘*time_from_infection*’ by ‘*grouped_severity*’.

All P values were adjusted using the Benjamini-Hochberg procedure ^47^.

### Cell type composition analysis - linear mixed effect model

The cell type abundances per sample were modelled using a generalised linear mixed model using a poisson outcome as described in Yoshida et al ^11^. We fitted log_2_ transformed age, and random effect terms on biological sex and inferred ethnicity, to account for collinearity with features of interest. We also fitted a random effect term on the donor identifier to account for donor-to-donor variation but captured the paired effects between longitudinal samples from the same donor. To perform longitudinal analyses, we modelled weeks since onset of disease (onset of symptoms or positive test, whichever came first) as categorical features, and scaled the conditional distribution of fold change estimates to the pre-infection samples that were available, and the pre-infection standard deviation was multiplied by the standard deviation of each other timepoint factor level to account for the increased variance that is introduced by scaling.

### Genetic principal component analysis

To overcome missing self-reported ethnicity data for some donors, we used PCA on genotyping data to quantify and infer genetic ancestry. We took the *souporcell* cluster genotypes of all donors and converted them into a numerical matrix to perform PCA on using *FactoMineR V2.4* ^48^. We then mapped self-reported ethnicity onto the genetic PCA results. This revealed that principal component 1 separated individuals with self-reported ethnicities indicating African ancestry from individuals with other ethnicities, while principal component 2 separated individuals of self-reported Asian ancestry from those with self-reported European ancestry. To adjust for the potential confounding effects of ethnicity (since ethnicity is associated with higher risk of severe and fatal COVID-19), we included these 2 principal components (continuous variables) as covariates in all linear mixed models (**Table 1, Supp.** Fig. 6).

### Differential abundance testing - steroid treatment

To examine the effect of steroid treatment on the cell abundance, MiloR package (v.0.99.0) ^23^ was used. The monocyte population was sub-setted to include only the samples from COVID- 19 positive patients who received the steroid treatment during ‘Wave 2’ of COVID-19. A KNN graph was constructed using the function ‘*buildGraph*’ (*k=30, d=30*) and the cells were assigned to the neighbourhoods on the KNN graph using the function ‘*makeNhoods*’ (*prop=0.1, k=30, d=30*). The number of cells belonging to each sample in each neighbourhood was counted using the function ‘*countCells*’. We included ‘*time_from_infection*’ in the design to account for the length of disease. SpatialFDR < 0.1 was used as a cut off point for significant enrichment/depletion.

### BCR and TCR data processing

Single-cell BCR and TCR data were initially processed with cellranger-vdj (v.6.0.0). Single cell TCR data was then converted into a cell by TCR format using scirpy v1.10.1 ^49^. BCR contigs contained in all_contigs.fasta and all_contig_annotations.csv were then processed further using *dandelion*^50^ singularity container (v.0.2.4) (https://www.github.com/zktuong/dandelion). BCRs were then matched to cell barcodes with *dandelion*.

### TCR analysis

After quality control, we recovered 197,330 T cells with fully resolved T cell receptors from 61 donors and across 187 samples. We identified 127,670 unique TCR clones, defined by a unique combination of CDR3a, TRAV, TRAJ, CDR3B, TRBV, TRBJ and donor, at the amino acid level. Of these, 93,960 came from COVID-19 positive ESKD patients and thus could be analysed longitudinally over the course of infection. A total of 3,727 clones (4%) were captured at two or more time points during infection. We further excluded all clones present in pre-pandemic samples for analysis related to COVID-19, as these could not have expanded in response to SARS-CoV-2, and finally obtained 3,137 clones (3.3%) for longitudinal analysis. Clonal frequency within a sample was calculated as total number of clone copies per sample over the total number of T cells within the sample. To determine expansion, only clones that were sampled at two time points or more within the 0 to 30 days after a positive PCR nasal swab, and that were absent in the pre-COVID-19 samples, were used. An expansion was noted if the highest clone frequency measured before a specific day since positive swab (cutoff) was lower than the lowest frequency measured after that day. If the clone was not sampled either before or after the cutoff, the respective frequency was set to 0. The cutoff at day 10 was selected as being in agreement with timing of an adaptive immune response. For the more stringent definition of expansion as determined by a dual cutoff, the clone frequency had to show an increase at the first cutoff and a further increase at the second cutoff. This allowed the capture of a steeper increase of clonal frequency over time, at the cost of considering fewer total clones.

SARS-CoV-2 specific TCR-epitope pairs were queried from VDJDB. Samples from before the pandemic were used to establish a baseline of matches with the database. While a single-chain match with the database only indicates a putatively binding TCR, quantifying significant differences in these numbers across T cell populations gives insight into antigen-specificity. Matches with the database were quantified for expanding and non-expanding clones using bar charts, where the error bars show variation across individual COVID-19 patients, and significance was determined with a two-sided Mann-Whitney test. To determine which clones were expanding the most, expansion was determined as the mean clonal frequency after the cutoff day divided by the mean clonal frequency before and sorted in descending order.

Activated T cells were identified by applying the automatic cell type classifier Celltypist (1.2.0, model = COVID19_HumanChallenge_Blood) and sub-setting to activated T cells.

Cell2TCR (0.1) was used on the clones that showed expansion according to the above definition using days 2 and 10 as dual cutoffs, and to generate TCR motifs, while excluding TCR sequences of MAI T cells.

TCR analyses were carried out in Python (3.10.2) using pandas (1.4.2), numpy (1.21.6) and scanpy (1.9.1), and visualised with matplotlib (3.5.2) and seaborn (0.11.2), in particular seaborn’s lineplot to show clonal frequency evolution. Statistical tests were carried out using the scipy.stats module (1.8.1) and plotted with statannotations (0.5.0). The regression line and R2 value were determined with the seaborn’s regplot function.

**Supplementary Table 1:** Metadata from individuals sampled.

**Supplementary Table 2:** List of antibodies included in the Total-seq panel.

**Supplementary Table 3:** Statistical tests from cell abundance analysis of cases versus controls.

**Supplementary Table 4:** Differentially expressed genes for each cell type for cases versus controls.

**Supplementary Table 5:** Gene set enrichment pathway analysis for each cell type based on DEGs from Supp. Table 4.

**Supplementary Table 6:** Statistical tests from cell abundance analysis of severity.

**Supplementary Table 7:** Differentially expressed genes for each cell type for severity.

**Supplementary Table 8:** Gene set enrichment pathway analysis for each cell type based on DEGs from Supp. Table 7.

**Supplementary Table 9:** Gene set enrichment pathway analysis of time x severity interactions.

---

**Supplementary Figure 1.**
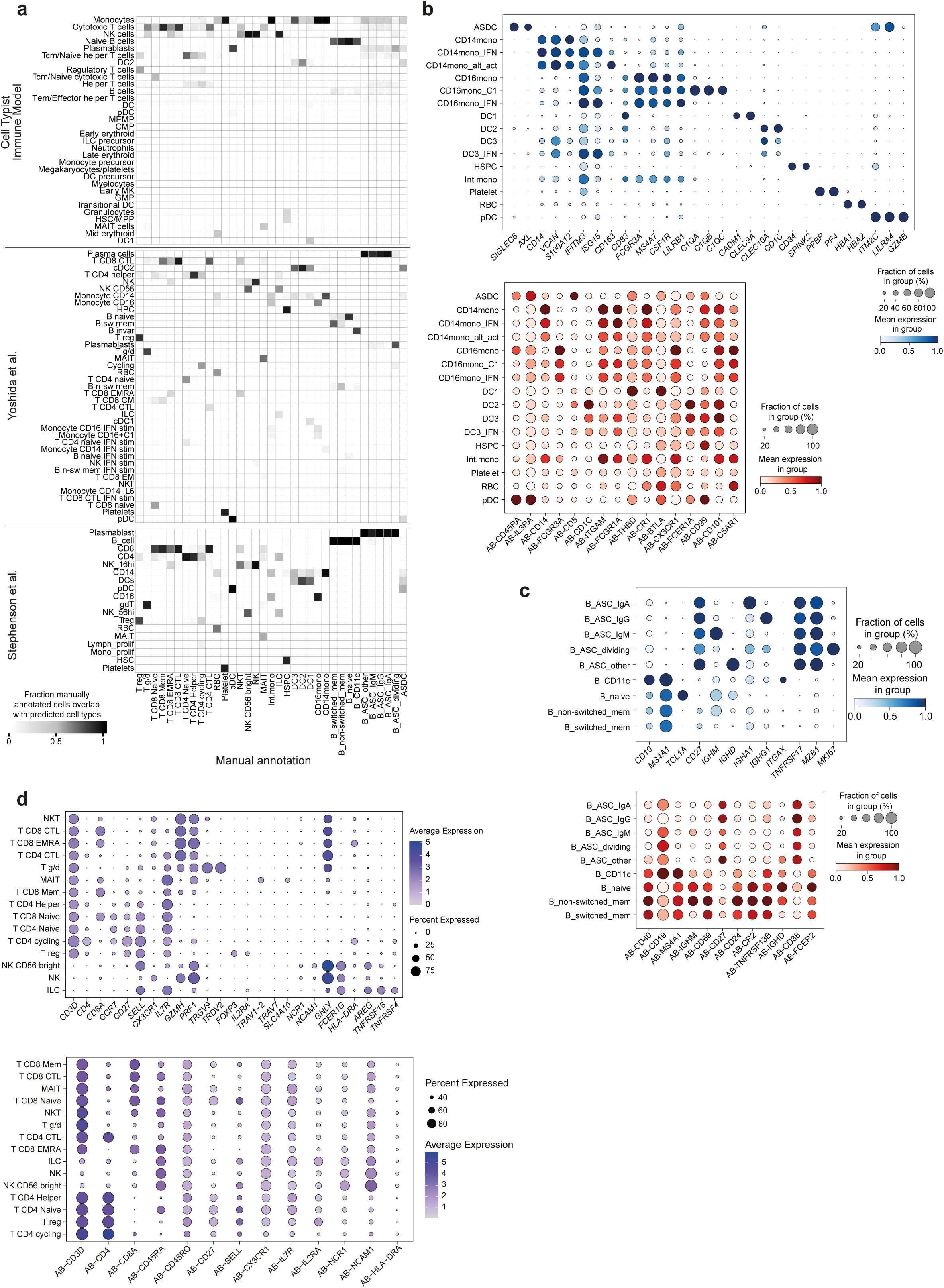
Cell demultiplexing and annotation. A. Heat map showing the overlap of manual versus predicted cell annotations. B. Dot plots displaying gene (top) and protein (bottom) expression of markers for myeloid and haematopoietic cells. C. Dot plots displaying gene (top) and protein (bottom) expression of markers for B cells. D. Dot plots displaying gene (top) and protein (bottom) expression of markers for T and innate lymphoid cells.

**Supplementary Figure 2.**
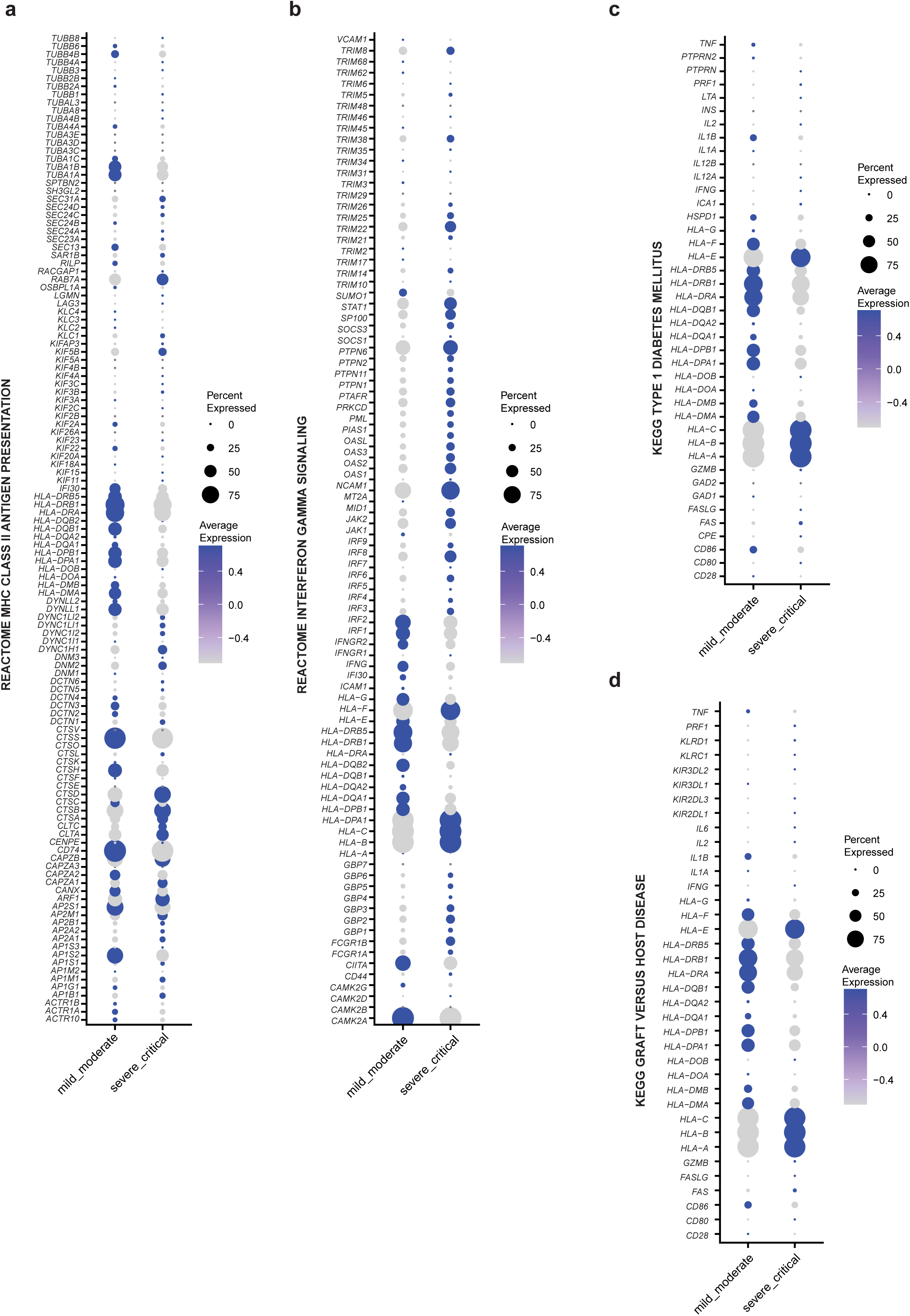
Expression of genes contributing to pathways in CD14 monocytes and B cells. A. Dot plot displaying the expression of genes that contribute to the Kegg ‘Systemic Lupus Erythematosus” pathway in CD14 monocytes. B. Dot plot displaying the expression of genes that contribute to the Kegg ‘Type 1 Diabetes Mellitus” pathway in CD14 monocytes. C. Dot plot displaying the expression of genes that contribute to the Kegg ‘Asthma” pathway in CD14 monocytes. D. Dot plot displaying the expression of genes that contribute to the Kegg ‘Graft Versus Host Disease” pathway in CD14 monocytes. E. Dot plot displaying the expression of genes that contribute to the Kegg ‘Allograft rejection” pathway in CD14 monocytes. F. Dot plot displaying the expression of genes that contribute to the Kegg ‘Allograft rejection” pathway in B cells.

**Supplementary Figure 3.**
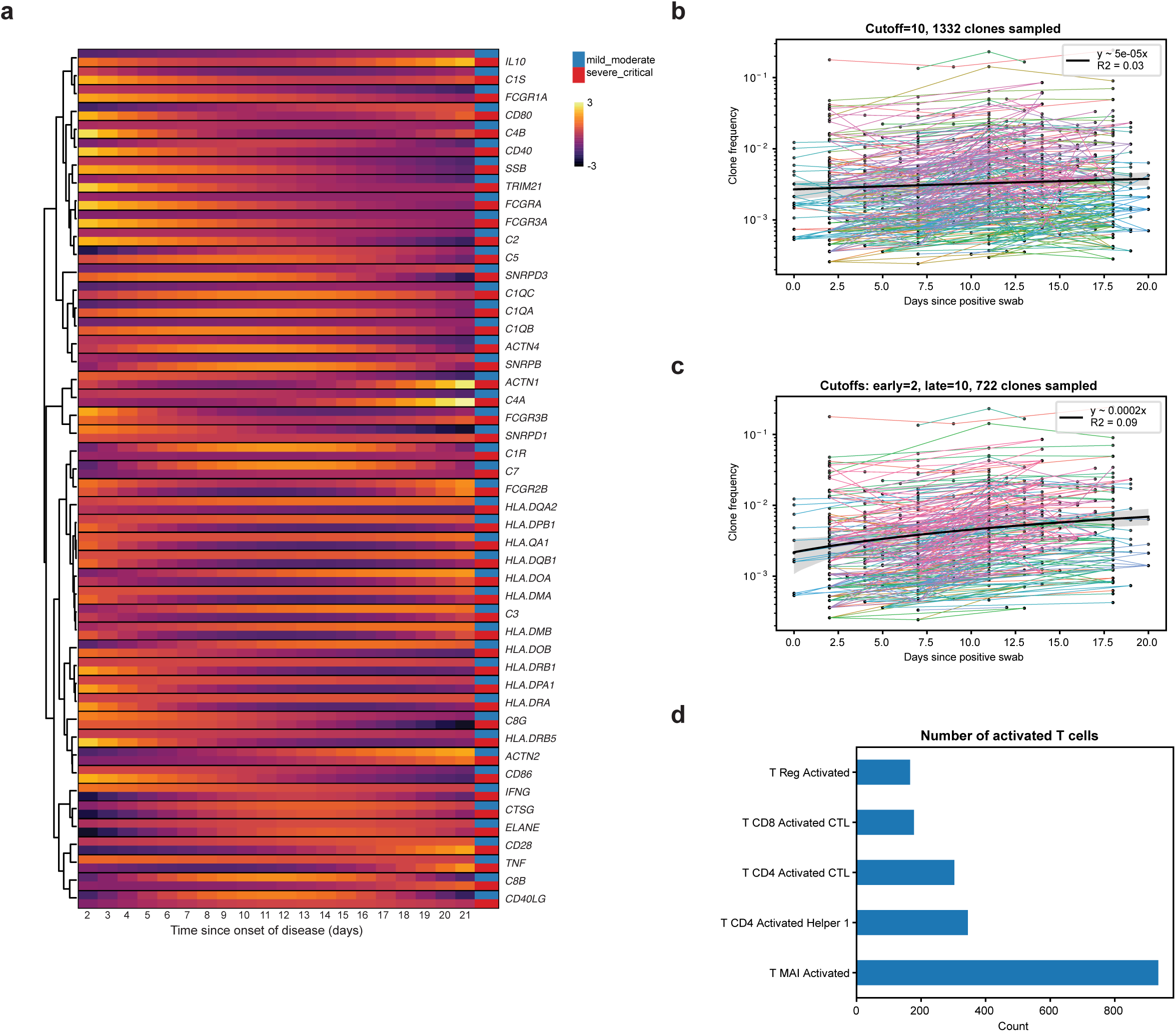
Longitudinal analysis. A. Clonal frequency dynamics for all clones expanded after day 2 post positive PCR result, as well as a trendline. B. Clonal frequency dynamics for all clones expanded after day 2 post positive PCR result and further expanded after day 10, as well as a trendline. C. Number of activated T cells according to Celltypist predictions, split by T cell type.

**Supplementary Figure 4.**
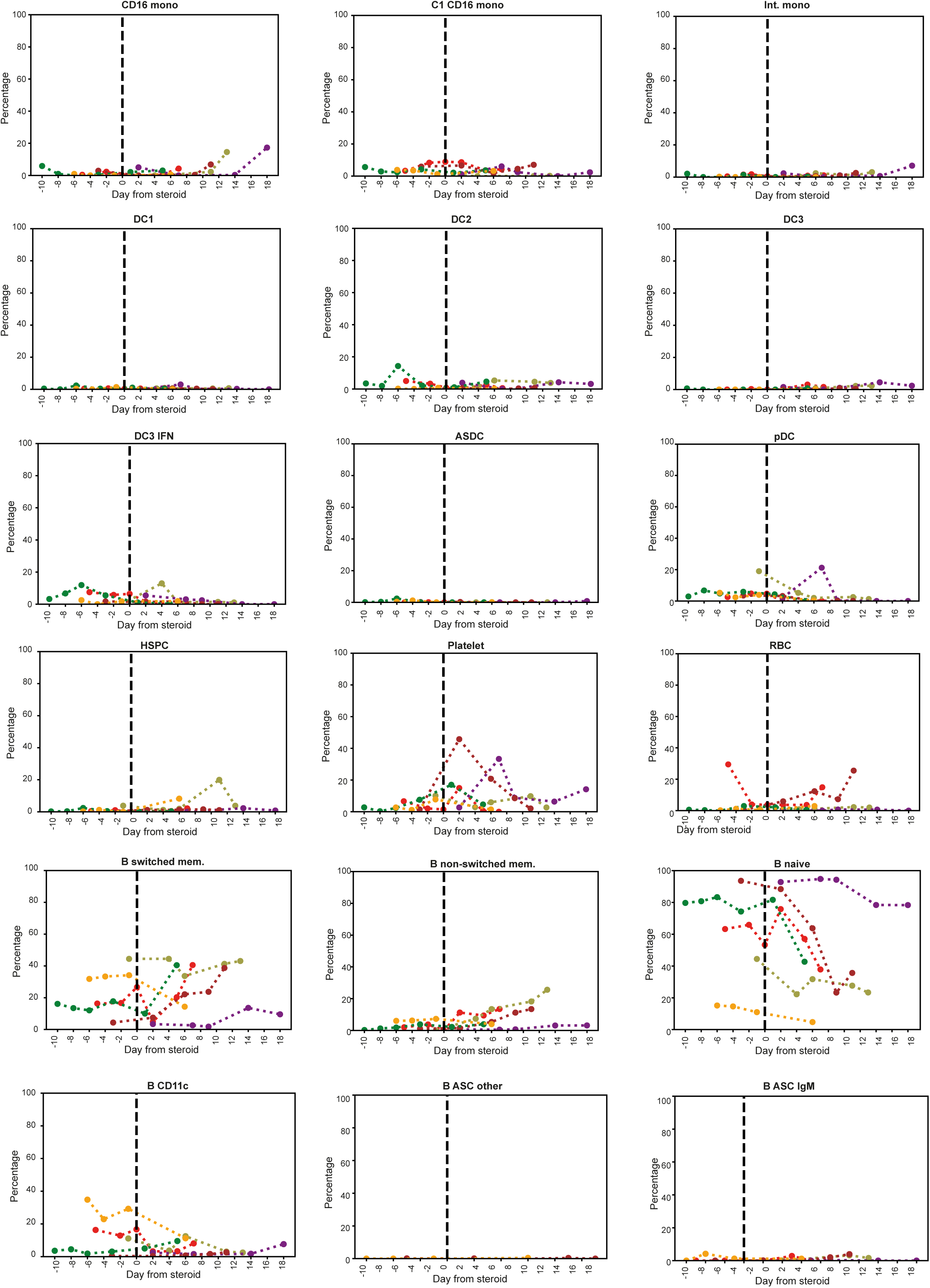
Trend of cell proportions during steroid treatment. Line charts displaying the percentage of cell subsets across the days before and after administration of steroids. Line colours represent different patients.

**Supplementary Figure 5.**
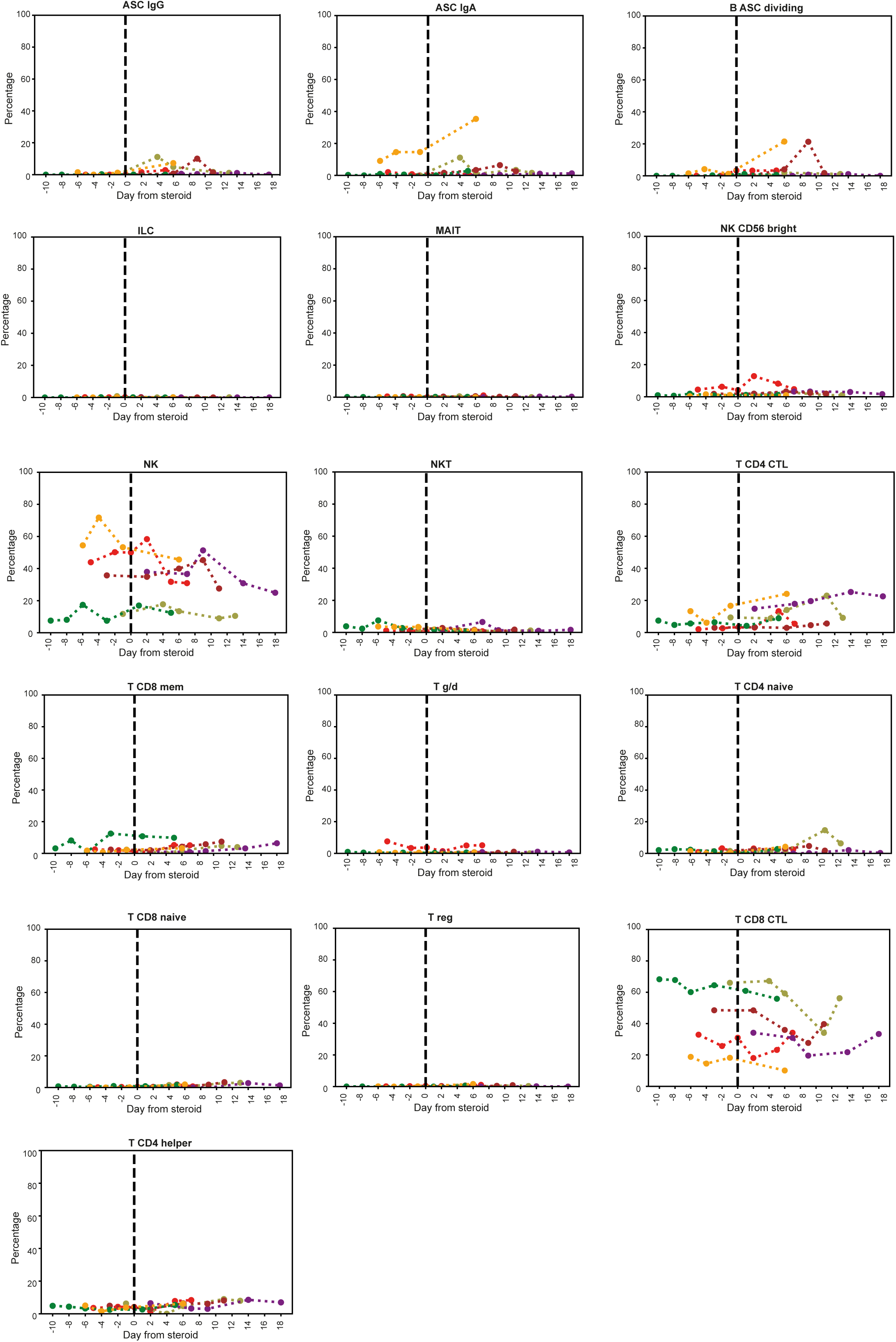
Trend of cell proportions during steroid treatment Line charts displaying the percentage of cell subsets across the days before and after administration of steroids. Line colours represent different patients.

**Supplementary Figure 6.**
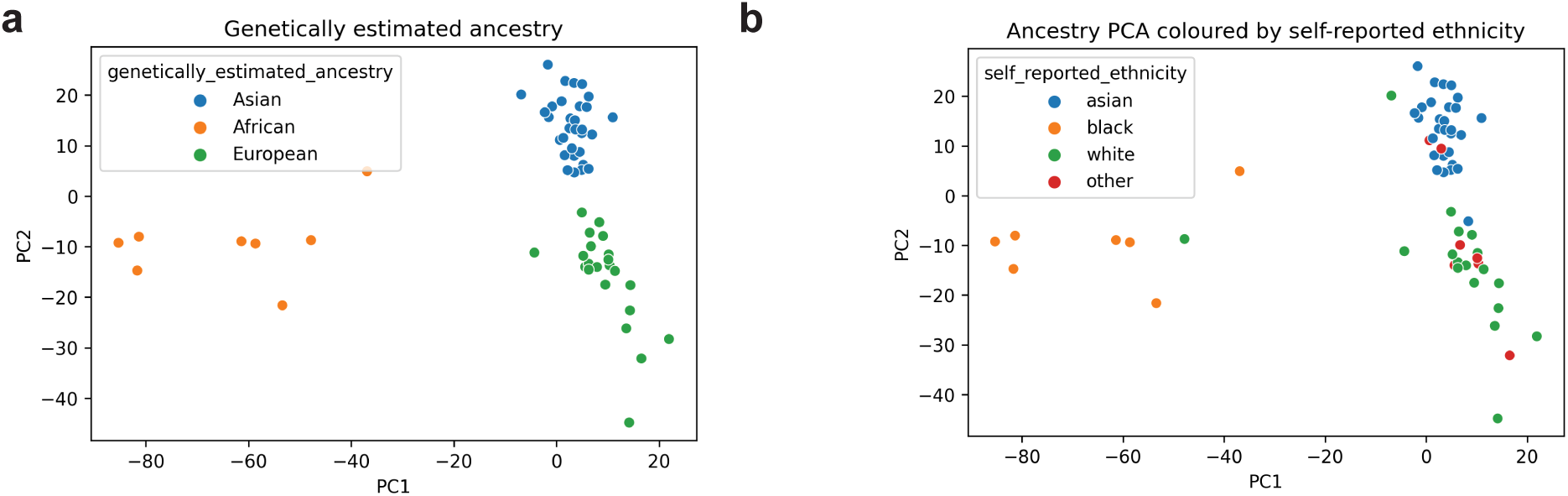
Estimated ancestry calculated using genotype compared to self-reported by patient. A. PCA plot of genetically estimated ancestry of each patient calculated using genotypes. B. PCA plot of the ancestry reported by the patients themselves.

